# The Population-based Microbiome Research Core: a longitudinal infrastructure for assessment of household microbiome and human health research

**DOI:** 10.1101/2021.11.22.21266369

**Authors:** Amy A Schultz, Kristen MC Malecki, Elizabeth A Holzhausen, Pravleen Bajwa, Paul Peppard, Tamara LeCaire, Shoshannah Eggers, Nasia Safdar, Ajay K Sethi

## Abstract

**Purpose:** The Population-based Microbiome Research Core (PMRC) is an expandable and longitudinal research core infrastructure to support the study of the human microbiome within the context of environmental, sociodemographic, and health factors. Broadly, the purpose of this infrastructure is to provide new insights into how human-environment interactions affect health through its influence on the composition and function of the microbiome. The PMRC was established as an ancillary study of the Survey of Health of Wisconsin (SHOW) and serves as a platform for ancillary studies, ongoing follow-up of the cohort, and expansion of the microbiome biorepository.

**Participants:** The study recruited adult participants who had previously participated in SHOW’s Wisconsin Microbiome Study (WMS). Over 59% of the eligible WMS participants agreed to provide a repeat stool sample and household samples including dust, high touch surface swabs and outside soil.

**Findings to date:** PMRC includes 323 individuals; the majority (96%) were over the age of eighteen, white (84%), urban (75%), and lived in their homes for over one year (92%). Overall, 97% of participants completed the questionnaire and household high-touch surface swab collection, and 93% and 94% completed dust and stool collection, respectively. Soil samples were collected for 86% of all participant homes.

**Future plans:** Sample protocols developed for the PMRC offer a unique framework for future household-based microbiome research. This infrastructure can support the generation of new knowledge on the role of the home environment in relation to the human microbiome and identify new opportunities for intervention research.

## INTRODUCTION

Multi-drug resistant organisms (MDROs) pose a significant threat to population health and have been identified by the World Health Organization as a global health priority. Yet, few population-based microbiome studies collect samples necessary to understand how microbial content in people’s homes contributes to gut microbial composition, function, and potential for increased MDRO colonization. Home environments are an important source of exposure and spread for many bacterial and other xenobiotics which collect on dust and on surfaces in homes ^1,2,3^. The contribution of household high-touch surfaces and household dust in shaping individual gut microbiomes is largely understudied. A better understanding of the microbiome interactions between humans and the environment requires more holistic approaches to data collection which include not only human and environmental specimen across a diverse geographic population over time. The Population-based Microbiome Research Core (PMRC) was established as a longitudinal and expandable biorepository for microbiome research that includes samples from multiple household media as well as individual level biomarker and health data.

The human gastrointestinal tract contains trillions of microbes, collectively known as the gut microbiota, which are key to many aspects of human health including immune function, nutrient and drug metabolism, and neurobehavioral traits.^4,5^ Imbalance or dysbiosis of the gut microbiome - the collective genomes of gut bacteria, viruses and fungi - is associated with several adverse health outcomes including obesity, risk of infection, inflammatory bowel syndrome, diabetes, allergic disease, and mental health conditions^5,6^. The gut microbiome is known to be influenced by genetics, exercise, diet, antibiotic use, and the environment^7,8,9^. Several studies have not only found the human microbiome can be affected by environmental factors, but that it can modulate responses to environmental factors through effects on metabolic and immune function^7,9-11^.

The last decade has seen a rapid expansion of research regarding human health and the gut microbiome. The White House Office of Science and Technology Policy announced the National Microbiome Initiative with investments of $121 million over fiscal years 2016 and 2017 and academic institutions have implemented initiatives including strengthening undergraduate curricula, advancing translational research, and expanding computational resources^10,11^. Expanding interest in the human microbiome has also resulted in several groundbreaking studies, including The Human Microbiome Project and The American Gut Project^11-13^. However, to our knowledge, no previous U.S.-based studies have established a longitudinal, population-based microbiome study that includes both human biomonitoring of gut microbiome and multiple household media.

While many studies have investigated the development of infant and child gut microbiomes over time, relatively less is known about longitudinal changes to the adult microbiome^14-17^. Few previous studies have investigated how the adult microbiome changes and adapts, and those that have found significant variability in microbiomes over time, with greater diversity being associated with more stability despite these studies’ limited sample sizes^18-20^.

The PMRC, an expansion of the Wisconsin Microbiome Study (WMS), builds on an unique opportunity to advance microbiome and health research to include individual microbiome and the contextual samples of indoor dust, surfaces and outdoor soil as predictors and sources of human microbial diversity. Building on previous microbiome studies, the PMRC is a relatively large, well-characterized, longitudinal study of adults with gut microbiome and household environmental samples. Furthermore, the PMRC utilizes the Survey of the Health of Wisconsin (SHOW)^21^ cohort and can leverage participant health data including information about diet and food insecurity, health and healthcare, insurance, medication use, mental health data, home environment and housing characteristics, and physical measurements such as blood pressure and body mass index. Throughout the design and implementation of PMRC it became clear there were no comprehensive, standardized protocols for home microbiome sample collection in soil, dust or surfaces exist. The following describes the origins of the PMRC resource, study population, and key findings to date.

## STUDY DESCRIPTION

### Overview

The PMRC leverages the existing Survey of the Health of Wisconsin (SHOW), a population-based health examination survey established in 2008. Details of SHOW data collection have been previously described^21,22^. In brief, the core SHOW study includes a randomly selected statewide population-based sample on over 5,000 state residents. A wide range of health, behavior, and environment data along with biometrics and biological specimens have been collected. In 2016, the Wisconsin Microbiome Study (WMS) was initiated as an ancillary study to expand the SHOW core survey to include microbiome data collection and relevant dietary assessments, questions, and risk factors. Details regarding the WMS have also been previously described^23^.

The PMRC was designed with the goals of characterizing home environment microbiome samples and longitudinal changes in human microbiome to increase understanding of home and outdoor environmental factors with individual gut microbial diversity and anti-microbial resistance in the population. Protocols included repeated stool samples and first-time collection of environmental samples, including outdoor surface soil, indoor settled dust, and indoor high-touch surface swabs. All samples were collected and banked for future research. Table 1 outlines the various study components. The study consisted of two in-home visits by a SHOW field staff member: (1) the in-home recruitment visit, where the participant consented, scheduled the in-home sample collection visit, and received a self-administered questionnaire and stool collection kit with instructions, and (2) the in-home sample collection visit where a SHOW field staff member collected the self-administered questionnaire, stool sample, and dust, surface swab, and soils samples. At the in-home recruitment visit participants were asked not to clean their home 24 hours prior to the in-home sample collection visit to ensure there would be enough dust to sample.

**Table 1.**
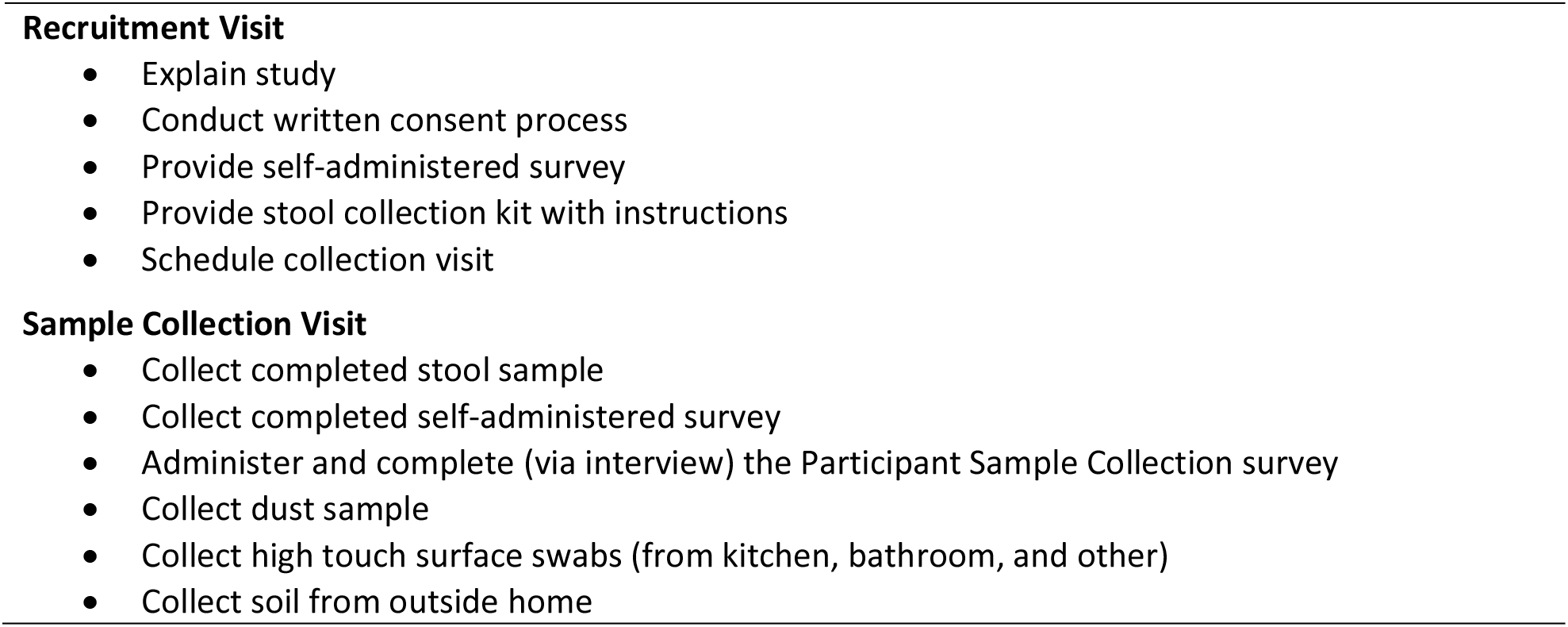
Components of the PMRC study, as conducted by the trained field staff member. Both visits are completed at the participant’s home.

### Recruitment and compensation

Subjects were eligible to participate in the PMRC if they were WMS participants, had provided a viable stool sample and completed a detailed dietary questionnaire during the initial WMS. A total of 543 (76%) of the original 713 eligible participants with baseline stool and diet data were contacted for recruitment into the follow-up study. Participants were invited based on feasibility and if they lived in study locations where resources were available to support follow-up sample collection. They were selected to have equal representation to overall urban and rural demographics compared to all potential eligible WMS participants. Lastly, the selected household had to be the individual’s usual place of residence, and if the individual had two residences, they had to spend a greater number of nights at the selected residence. All participants completed an informed consent at the in-home recruitment visit, as approved by the University of Wisconsin-Madison Institutional Review Board. More than one individual per household could be eligible to participate. Participants were monetarily compensated for completing each part of the study.

### Questionnaires

A self-administered survey questionnaire was given to every participant at the in-home recruitment visit and collected at the in-home sample collection visit. The questionnaire consisted of personal health history and health history of other household members pertaining to appendectomy, surgery, medical devices, dialysis, infection, antibiotic/probiotic use, and time spent in a nursing home, hospital or inpatient rehab facility in the last 12 months. These questions were chosen as important factors which can affect pathogenic and non-pathogenic components of the microbiome or are important risk factors of MDRO. Additional questions were taken from the SHOW survey questionnaire which may change over time and act as important confounders, such as smoking and physical activity.

The questionnaire additionally asked about household ventilation, heating/cooling, cooking appliances, dehumidifiers, leaks, and potential environmental exposures near the home. Habitual questions were also asked about cleaning and cleaning products; water use, filtration, and treatment; bathing and showering; and time spent in and outside the home. The household questions were selected to provide data on the important factors which can change the environmental conditions and ultimately affect the microbiome in the air, dust, and surfaces within the home. The habitual questions offer insight into potential windows of environmental exposure. The household and habitual questions were selected from the main SHOW survey or were borrowed from the Home Environment Questionnaire asked in the Canadian Child Health Study^24^. Complete questionnaires can be found in the online supplement 1.

Additional interview questions asked during visit two pertained to cleaning and cleaning products used within the last 24 hours. These questions were asked at the household level and not the individual level. If more than one person participated per household, only one volunteered to answer the household level questions.

### Environmental and Biological sampling

All environmental samples were collected by trained field staff following strict protocols designed to ensure high quality, reproducible samples and to avoid contamination. Field staff wore a new pair of non-latex gloves when performing each environmental sample collection. Dust was collected at the individual level, resulting in a dust sample for each participant. Soil and high-touch surface swabs were collected at the household level, meaning if more than one individual participated per household, only one set of high-touch surface swabs and soil were collected.

#### Dust

Dust was collected with a Sanitaire® canister vacuum (Model #SC3683B) fitted with a DUSTREAM™ collector (Indoor Biotechnologies, Charlottesville, Virginia, USA). Participants self-reported the most used room during waking hours and identified the most frequently used chair in this room, and the chair’s surface was vacuumed continuously for three minutes, split evenly between the back and lower parts of the chair. Using the same DUSTREAM™ collector, a one-meter squared section of the floor adjacent to the chair was also vacuumed continuously for two minutes. After collection, the DUSTREAM™ canister was fastened closed, placed in a sterile plastic bag, and immediately placed in a cooler with ice packs. SHOW field staff wore disposable sterile shoe covers and kept equipment away from the sampling area to avoid contamination.

#### Soil

Soil samples were collected from up to three areas in participants’ yards or outdoor spaces near their residence. One soil sample was taken as close as possible to the entrance/exit of the home used by the participant most often. The second soil sample was taken from an outdoor location the participant or other household members spent the most time near (i.e. walkway to mailbox or car, near patio, grill, or flowerbed, etc.). The third soil sample was taken from a produce garden, only if participants or other household members spent time in the produce garden during the growing season. Soil samples were not collected from participants reported that they did not have access to a yard or outdoor space.

A soil corer and a metal lab scoopula or spatula were used to collect outdoor soil samples from each household. First soil was cored and removed to “cleanse” the core of soil from prior participants’ homes before coring for sample collection. Next, the corer was used to break up the surface soil again and the corer was sunk straight down to gather dirt 6-12 inches below the surface, avoiding any roots and the topsoil layer. Each sample was transferred to a maximum of three one mL cryovials of soil from each location within the yard. Immediately after collection and processing, soil samples were put in sterile plastic bags and placed in a cooler with ice packs. Soil corers, metal scoopulas and spatulas were cleaned with water between each of the three sample collection areas in yards.

#### High-touch Surfaces

High-touch surface samples were collected using regular, flocked ESwabTM (COPAN Italia S.P.A., Brescia, Italy). The kit includes a swab stick and liquid based multipurpose collection and transport system that maintains the viability of aerobic, anaerobic and fastidious bacteria for up to 48 hours. A total of three swabs were collected in each household by trained SHOW field staff; one swab was collected in the kitchen, one in the most frequently used bathroom, and one from frequently used surfaces in the home. All rooms and items sampled were identified by the participant.

For the kitchen swab, the most frequently used cutting board, refrigerator produce drawer, the refrigerator handles, and an area of the kitchen counter were each swabbed for 15 seconds using the same swab prior to placement in the collection tubes. The bathroom collection consisted of swabbing the following items for 15 seconds each: the sink handles, the handle on the inside of the door, the bathroom counter, and the toilet lever. For the final swab, participants identified two doorknobs frequently used and two additional surfaces touched frequently (e.g. cell phone, TV remote, light switch etc.). The two doorknobs and the two other surfaces were swabbed for 15 seconds each and then the swabs were placed in collection tubes. Field staff wore gloves and followed protocols to avoid sample contamination during collection. After the three samples were collected, each collection tube was placed in sterile plastic bags and immediately stored in a cooler with ice packs.

#### Stool

Participants self-collected a stool sample at home using a sample collection kit provided during first recruitment visit. The collection kit included a sterile 60 mL specimen cup, a sterile wood tongue depressor, gloves, a specimen label, a biohazard bag, a brown paper bag and an instruction sheet. Participants were instructed to collect the sample within 24 hours of their scheduled home visit and refrigerate the sample until it was picked up by the field interviewer during visit two.

### DESCRIPTIVE CHARACTERISTICS

Overall, 323 (59%) of those invited agreed to participate, completed the follow-up questionnaire, and contributed at least one biological sample. A comparison of the demographic characteristics of the PMRC participants with original WMS parent study can be found in Table 2. PMRC participants were slightly more likely to be 60+ years of age and female when compared to those who participated in the WMS. We did not find statistically significant differences in urbanicity, home ownership, or residential mobility in the last year among participants in PMRC and WMS.

**Table 2.** Demographic characteristics of Wisconsin Microbiome Study (WMS) participants who were eligible for the Population-based Microbiome Research Core (PMRC), and those who agreed to participate in PMRC.

| Characteristic |  | WMS<br>n = 543<br>n (%) | PMRC<br>n = 323<br>n (%) | P-trend<sup>a</sup> |
| --- | --- | --- | --- | --- |
| Age (years) |  |  |  | 0.007 |
|  | 18-29 | 49 (9.0) | 14 (4.3) |  |
|  | 30-59 | 251 (46.2) | 137 (42.4) |  |
|  | 60+ | 243 (44.8) | 172 (53.3) |  |
| Gender |  |  |  | 0.59 |
|  | Female | 311 (57.3) | 191 (59.1) |  |
|  | Male | 232 (42.7) | 132 (40.9) |  |
| Race/ethnicity<br>(missing = 1) |  |  |  | 0.10 |
|  | White | 447 (82.5) | 273 (84.8) |  |
|  | Black | 57 (10.5) | 39 (12.1) |  |
|  | Hispanic | 20 (3.7) | 5 (1.6) |  |
|  | Other | 18 (3.3) | 5 (1.6) |  |
| Urbanicity |  |  |  | 0.98 |
|  | Urban | 408 (75.1) | 243 (75.2) |  |
|  | Rural | 135 (24.9) | 80 (24.8) |  |
| Home ownership<br>(missing = 1) |  |  |  | 0.60 |
|  | Own | 398 (73.6) | 244 (75.8) |  |
|  | Rent | 137 (25.3) | 73 (22.7) |  |
|  | Occupied (no rent) | 6 (1.1) | 5 (1.6) |  |
| Residency duration<br>(missing = 18) |  |  |  | 0.74 |
|  | <=1 year | 42 (8.0) | 27 (8.7) |  |
|  | >1 year | 483 (92.0) | 285 (91.4) |  |

<sup>a</sup> p-trend: statistical significance by Chi-square test

Table 3 displays completion of each part of the follow-up study. Ninety-seven percent of participants competed the questionnaire and high-touch surface swab collection. Ninety-three and ninety-four percent completed dust and stool collection, respectively. Soil had the lowest completion rate, of 87%, in large part due to widespread flooding across the state in the late fall of 2017 which limited the ability to collect soil samples in some areas.

**Table 3.** Completion (%) of each part of the PMRC study

| Study component | N (%) |
| --- | --- |
|  | **Individuals (n = 323)** |
| Questionnaire | 314 (97.2) |
| Stool sample | 304 (94.1) |
| Dust sample | 301 (93.2) |
|  | **Households (n = 230)** |
| High touch surface swabs | 224 (97.4) |
| Soil samples from yard | 199 (86.5) |

### The PMRC as a platform for longitudinal microbiome research

One of the main goals of the PMRC is to serve as a platform for ancillary studies, ongoing follow-up of the cohort and expansion of the biorepository for integrated microbiome research. Several studies are ongoing, including (1) translational research using existing gut and oral microbiome data to test new and effective antimicrobial drugs and therapies to combat emerging resistant pathogens, (2) expansion of the biorepository and cohort to include children in order to assess the microbiome at several body sits and MDRO colonization in children attending day care, (3) expansion of the biorepository to include PBMC blood sample collection in order to determine how gut microbial composition and the resulting co-metabolome varies with chronological age and biological aging rate, (4) use of the existing data and biorepository to determine how gut dysbiosis and diversity is affected by heavy metal exposure and associations with metabolism, homeostasis, and inflammation. PMRC samples can expand the utility of these studies to understand household and outdoor contributions to human microbiome diversity, composition and function. For more information on using these data or leveraging the Wisconsin Microbiome Study or SHOW for research grants or studies, contact the study investigators or visit the SHOW website: https://show.wisc.edu/.

## STRENGTHS AND LIMITATIONS

The PMRC project has determined that field sample collection for microbiome samples from multiple sources inside and outside the home, and from individuals, is feasible using standard protocols and trained field interviewers. The PMRC is one of the first population-based microbiome research platforms designed to collect data within a geographically diverse urban, rural, and suburban population. As an ancillary study to the SHOW program, the PMRC provides a rich data source with phenotypic variety. The PMRC enables examination of the microbiome relationship between humans, their household micro-environment and the animals inhabiting it. As such, the PMRC also recognizes the importance of holistic approaches to understanding determinants of human microbial diversity that may offer new insights for treatment and interventions aimed at modifying microbial composition, with an effort to improve health.

Other previous human microbiome studies and cohorts have been conducted, but they have relatively smaller sample sizes compared to the WMS and the PMRC and they do not include corresponding sample collection of household dust, surface or outdoor soil. The Human Microbiome Project was a clinical study, sampling 300 healthy individuals in Houston, Texas and St. Louis, Missouri,^11^ offering limited generalizability with respect to collecting samples in both urban and rural areas. The American Gut Project (AGP), based at the University of California-Davis is larger, but relies on volunteers to be citizen scientists who self-collect biospecimens and send them in for analysis, with limited characterization of individual vectors such as frequently used household surfaces and soil as potential sources of microbial diversity in the population^12^. While crowdsourcing and citizen science offer a novel opportunity for participants and researchers, this model may result in a biased cohort. Further, participants in the AGP were six times more likely to report an inflammatory bowel disease diagnosis compared with the US population^13^. In contrast, the PMRC participants provide a wealth of data on social determinants of health, health behaviors, and existing health status in addition to two stool microbiome samples in over 300 individuals. Furthermore, the additional environmental samples can be used to increase understanding of the role of micro-environments on gut microbial diversity, composition and function and support new microbiome-based prevention and interventions. Clinical research rarely considers these contextual drivers of microbiome and sources of bacterial exposure outside of hospital settings, thus the PMRC has a great deal of potential in advancing the field.

The PMRC mission is to provide an expandable and longitudinal human and environmental microbiome repository for investigators to carry out microbiome and multi-drug resistant research spanning translational, clinical, and epidemiological designs and serve as a resource for preliminary data for extramural grant submission. The collective use of the infrastructure aims to facilitate novel research, new public health interventions and clinical treatments, and move the microbiome science and knowledge forward.

The PMRC fills a critical gap in existing human health studies where a lack of well-characterized household environmental samples and longitudinal individual microbiome data exist. Humans likely acquire many of their bodily microorganisms through direct contact with their surroundings and tend to share more microbes with individuals and pets they are in frequent contact with. While several studies are beginning to characterize the microbiome of the indoor environment, not as many have linked these characterizations to the human microbiome and human health^25-32^. Additionally, many environmental-human microbiome studies occur in isolation and do not span the same study sample and use the same data collection and processing techniques, making comparisons across studies difficult^33,34^. Ensuring findings are consistent across and within study samples is an important next step in microbiome research^35^. Furthermore, while baseline and follow-up stool samples exist for longitudinal analysis, environmental data is only cross-sectional and requires additional funding for analysis. However, with limited human-environmental microbiome research at present, and the ability to expand and follow-up the SHOW cohort, the repository presents a unique opportunity for preliminary research for pilot grants. While soil was collected on 87% of participants, the remaining survey, stool, and environmental samples were successfully collected on over 93% of participants. It is important to consider weather as a predictor of soil collection and time collection of field samples accordingly. Overall, the response rate for longitudinal follow-up and environmental sample collection has high, adding to the richness of the research platform and limiting selection bias due to loss of follow-up.

### Future Plans

Future directions of the Wisconsin Microbiome Study include analyzing changes in microbiome metrics longitudinally. In additional, plans are underway to characterize the indoor home microbiome and investigate how it may be related to human microbiome and human health. There are several avenues upon which investigators could analyze land use, outdoor environmental, and its relationship to environmental microbiome, human microbiome, and health. Our hope is the WI Microbiome study platform and cohort continue to grow, and new knowledge, treatments, and interventions are discovered as a result of the infrastructure and the gracious participants who have volunteered their time, data, and biosamples. We plan to continue to encourage use of the WI Microbiome study’s repository and seek collaboration with other interested researchers.

## Supporting information

Dust Collection Protocol

High Touch Surface Swab Collection Protocol

Participant Sample Collection Survey

Participant Self-Administered Questionnaire

Soil Collection Protocol

Stool Collection Instructions for Participant

## Data Availability

The PMRC encourages outside collaborations with University of Wisconsin-Madison researchers from other institutions and research organizations. Grants which expand on the WI Microbiome Study biorepository are encouraged. Researchers can apply for data, or analysis of biomaterial, by submitting a proposal to the Wisconsin Microbiome Study Scientific Board. All proposals will be reviewed on scientific contribution, quality, and methodology. For more information on using these data or leveraging the Wisconsin Microbiome Study or SHOW for ancillary studies, visit the SHOW website: https://show.wisc.edu/

## COLLABORATION

The PMRC encourages outside collaborations with University of Wisconsin-Madison researchers from other institutions and research organizations. Grants which expand on the WI Microbiome Study’s biorepository are encouraged. Researchers can apply for data, or analysis of biomaterial, by submitting a proposal to the Wisconsin Microbiome Study Scientific Board. All proposals will be reviewed on scientific contribution, quality, and methodology. For more information on using these data or leveraging the Wisconsin Microbiome Study or SHOW for ancillary studies, visit SHOW’s website: https://show.wisc.edu/.

