## Supplementary material for "The Population-based Microbiome Research Core: a longitudinal infrastructure for assessment of household microbiome and human health research": Dust Collection Protocol

#### 1.) Overview

You will collect a total of 1 dust sample from the participant's Most Lived-in Space

Sampling should occur in this order:

1. Chair/couch in most lived-in space,
2. Floor in most lived-in space

| MOST LIVED-IN SPACE DUST SAMPLE OUTLINE | |
|---|---|
| Location of vacuuming | Time spent vacuuming |
| **Most lived-in chair/sofa/couch, etc.**<br>*(if applicable and not wood/metal)* | **3 minutes** |
| • Vacuum seat cushions, seat back and arms of the chair. Try to split time evenly among the different parts to be vacuumed.<br>• If two pieces of furniture are used equally (within that room) then both pieces of furniture can be vacuumed for 1.5 minutes each.<br>• If most lived-in space does not have a chair (or chair is wood/metal), move on to next section. | |
| **Floor/carpet near most lived in chair/sofa** | **2 minutes**<br>*(or 5 Minutes, if no chair)* |
| • $1m^2$ area<br>• If it is not possible to collect a 1 m2 sample, sample the available area for 2 minutes. | |

#### 2.) Important things to remember:

1. Immediately upon entering the household, determine where the most-lived in space is. (You do NOT want to sit on the most lived in chair/sofa or walk on the floor near it while conducting other parts of the home visit until after sampling).

2. Do NOT vacuum stuffed animals. Rolled up blankets, towels etc. that act as pillows on a couch or chair should not be unrolled if vacuumed as a pillow.

3. If you need to move any items in the most lived-in room, be sure to put them back to their original location when you are done sampling.
4. A single clean Dustream dust collector will be used to sample the furniture and the floor. The furniture should be sampled before the floor because the same device will be used for the entire time.

5. For the most lived in chair/sofa, if the participant does not identify one, if the participant stands or lays on floor in most lived in space, or if participants most lived in chair cannot easily be vacuumed (wood / metal) , then vacuum floor for 5 minutes instead of 2.

#### 3.) List of Supplies:

a. Dust Kit
    i. Small Ziplock Bag:
        1. DUSTREAM collector nozzle with:
            a. Filter
            b. Two end Caps
    ii. 1 pair of disposable booties
b. Vacuum with power cord
c. Vacuum wand
d. Extension cord
e. 3-to-2 prong adaptor
f. 2 folding meter sticks
g. Timer
h. Extra AAA batteries
i. Non-latex gloves

### Dust Collection
### Step-by-Step Guide

#### Location: Most Lived-in Space

*There are accompanying questions in REDCap that will help you identify where to vacuum in the participant's home. Once you have identified the locations, use the following step-by-step guide to collect each of the samples. This guide walks you through sampling of the* ***participant's most lived-in space.***

**PHOTO CORRECTIONS:** You will have 2 meter sticks, and will NOT use a tape measure as shown in the photos below. Biohazard bags will not be used, Sample collection kits will use plain ziplock bags

| Instruction | Notes or pictures |
|---|---|
| **A. Prepare for sampling:**<br><br>**1. Identify where the participant's most-lived in space in the home is and where in this space he/she sits (or stands).**<br><br>**2. Transport your vacuum and dust sample collection supplies & materials to that room.** | *Use REDcap dust collection form questions to identify where the participant spends most of his/her waking hours when in the home.* |
| **3. Immediately upon entering the room, scan the room and locate the furniture and floor area you will sample.**<br><br>**DO NOT CONTAMINANT THE AREA WITH YOUR BELONGINGS or by WALKING IN THE AREA TO BE VACUUMED.** | 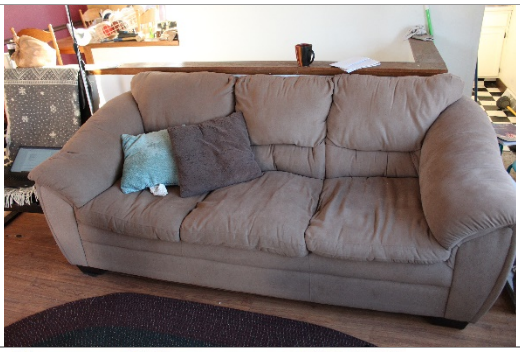 |
| **4. Remove a pair of booties from the Dust kit bag.**<br>**5. Put disposable booties on over your shoes/feet BEFORE approaching the chair/furniture and walking over the floor near where you will sample.** | 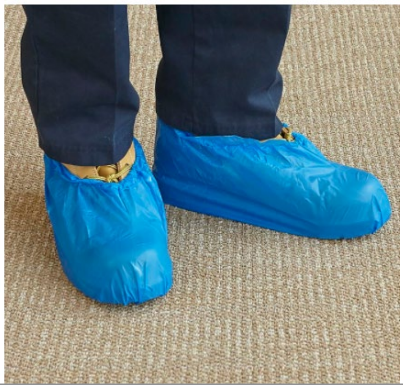 |

| | |
|---|---|
| **6. Remove the timer from your supplies and set out on a table, piece of furniture, or floor where you easily hear it beep and close enough as to not be inconvenient to reset.** | 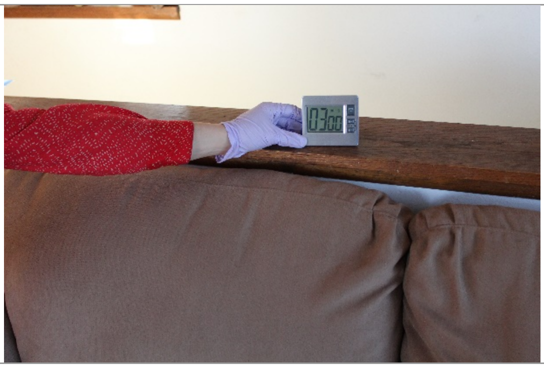 |
| ***7. Ask the participant if the use of the vacuum is likely to overload the circuits. If so, do not operate the vacuum in t he same room with a running air conditioner or electric heater. Ask participant to Turn off the air conditioner or heater before operating the vacuum. If a circuit does overload, assist the participant with their usual procedures.***<br><br>**8. Plug in the vacuum and make sure the cord will reach the area to be sampled.** | *Use an extension cord or 2 prong adapter if needed.*<br>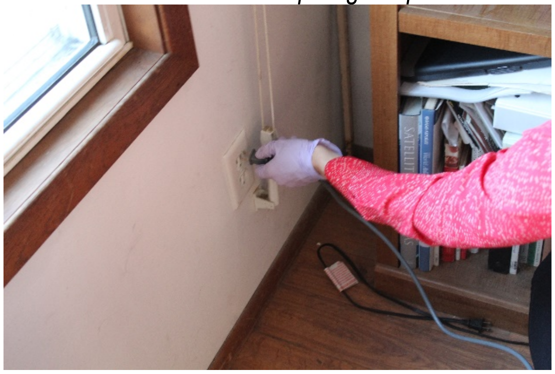<br>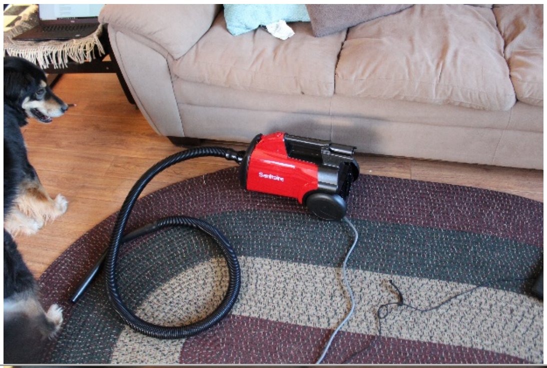 |
| **9. Put on a pair of disposable gloves. These should <u>always be discarded</u> <u>between rooms</u> and should not be reused. Dispose of gloves in YOUR garbage bag – <u>never in the participant's garbage</u>.** | 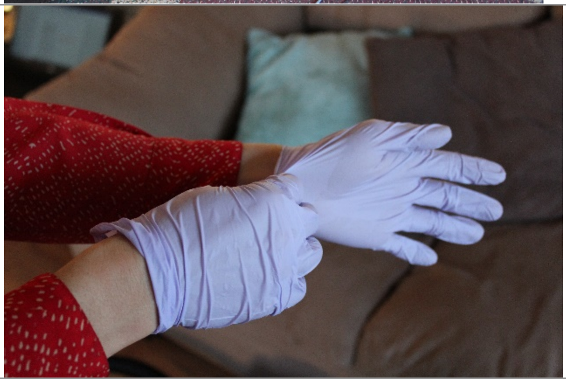 |
| **10. Open ziplock bag with unused filter**<br><br>**11. Open ziplock bag with Dustream collector and remove Dustream dust collector.** | *The only thing holding the dust filter in the nozzle is the suction of the vacuum. When the vacuum is NOT on, make sure the wand's nozzle end is facing UP so the filter and dust collected does not fall out!* |

12. **Grab the filter by the outside plastic casing and not the mesh and place it into the DUSTREAM collector.**

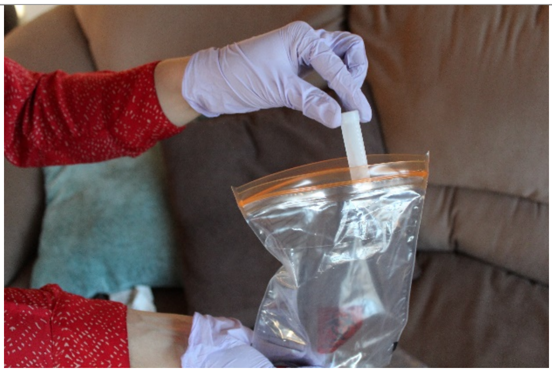

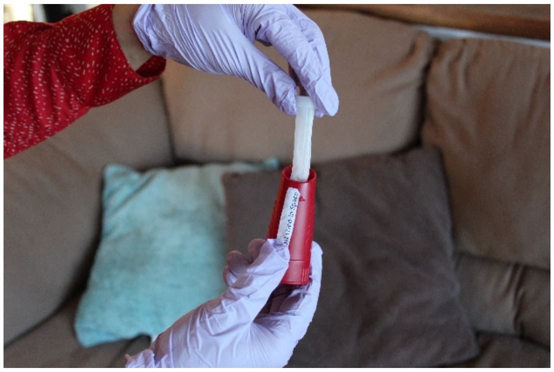

13. **Place the Dustream collector on the vacuum wand, holding the vacuum wand vertically, with the nozzle end facing up. The collector should already have a filter in it - ensure that it does.**

14. **Make sure the slot at the base of the wand is not exposed, but the sliding door is covering the window so that air does not escape.**

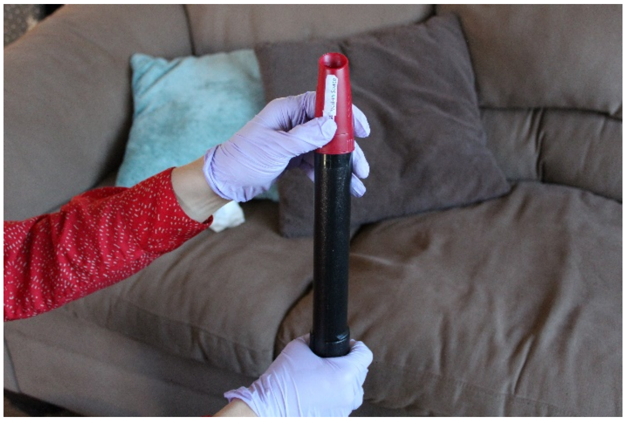

#### B. VACUUM the CHAIR the participants uses in the most lived in space

#### 3 minutes

**15. With the wand still upright, turn the vacuum on.**

**16. Set the time to 3 minutes.**

**17. Press start on the timer.**

**18. Vacuum the chair/couch for <u>3 minutes</u>:**

a. Vacuum seat cushions, seat back and arms of the chair. If the cushions are reversible, both sides should be vacuumed. Vacuum reversible cushions or any cushions that are not attached to the seat. Try to split time evenly among the different parts to be vacuumed.

b. If two pieces of furniture are used equally (within that room) then both pieces of furniture can be vacuumed for 1.5 minutes each.

c. If most lived-in space does not have a chair (or chair is wood/metal), move on to next section.

*IMPORTANT:*

*While vacuuming, angle the wand at about a 45 degree angle so the opening of the Dustream is in full contact with the surface to be vacuumed.*

*Slowly and firmly, rub the wand back and forth over the area to be vacuumed (1 foot per second).*

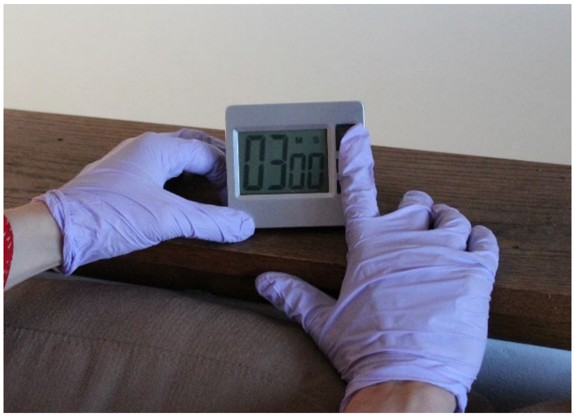

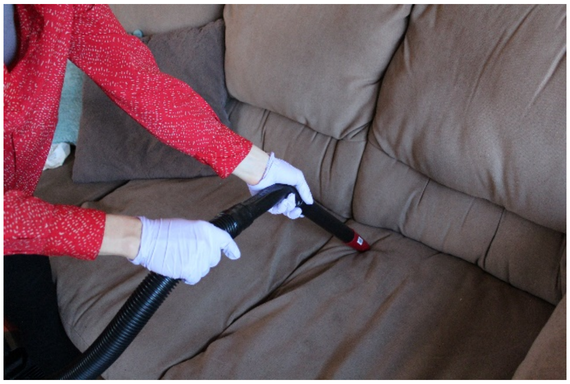

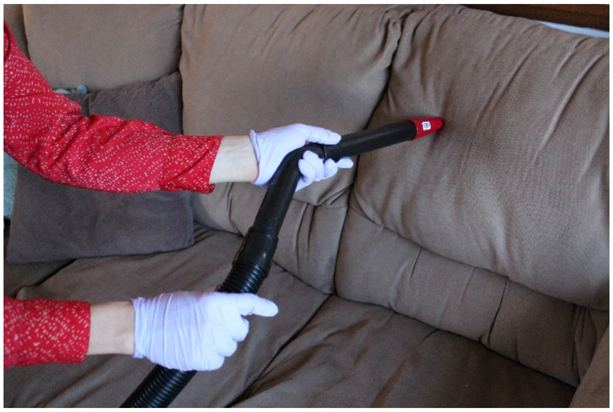

| | |
|---|---|
| | 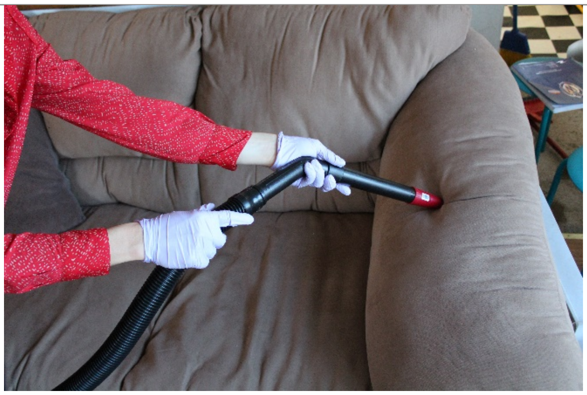<br>*Do not vacuum the area under the cushions or deep into crevices of the chair.* |
| **19. Stop alarm.**<br><br>**20. Holding wand upright, turn vacuum off.**<br><br>**21. Lean vacuum wand against vacuum, bed frame, or other piece of furniture while to prep for sampling the floor.** | 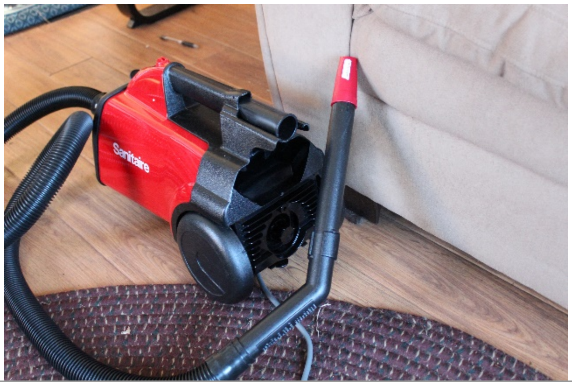<br> |
| ## C. VACUUM floor in most lived-in space where participant sits (or stands / spends most time)<br>## 2 minutes<br>**(unless did not sample chair, then 5 minutes)** | |
| **22. Place your two meter sticks in a L-shape right in front of (or next to) where the participant sits (or stands).** | 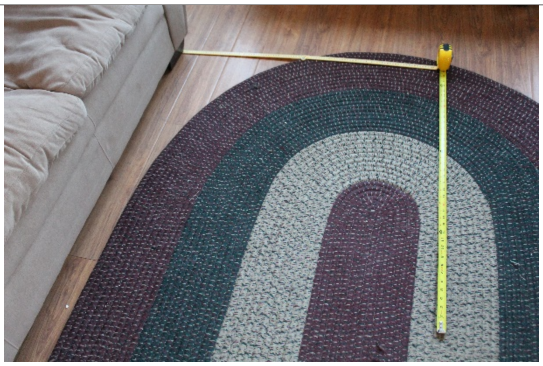 |

23. Set time to 2 minutes (or 5 if did not sample chair).

24. Press start on the timer.

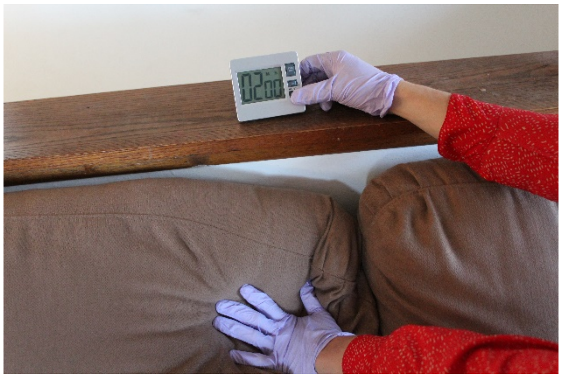

25. Vacuum a total of 1 $m^2$ of the living room floor for 2 minutes. If it is not possible to collect a 1 $m^2$ sample, sample the available area for 2 minutes.

    a. Vacuum in one direction for 1 minute.
    b. Rotate your vacuum strokes 90 degrees and continue vacuuming, covering the entire 1 $m^2$.

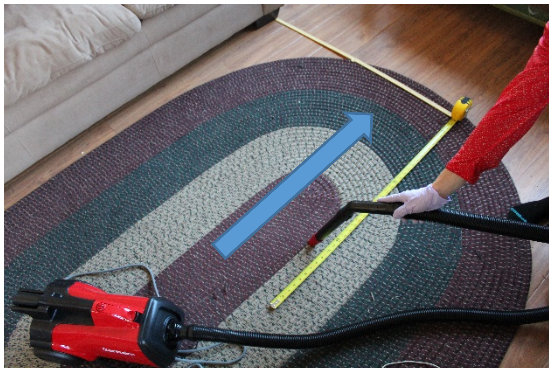

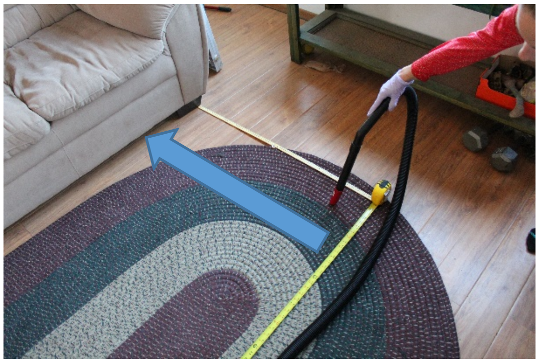

26. Stop alarm.

27. Holding wand upright, turn vacuum off.

28. Lean vacuum wand against vacuum, bed frame, or other piece of furniture while you prepare to vacuum the bedroom floor.

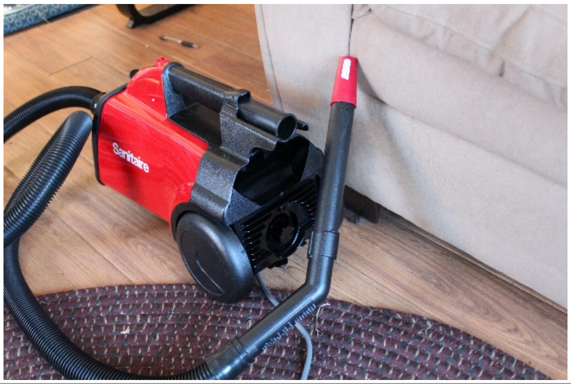

#### E. Finish and Cleanup

29. Remove the bag with the DUSTREAM nozzle caps from your kit bag.

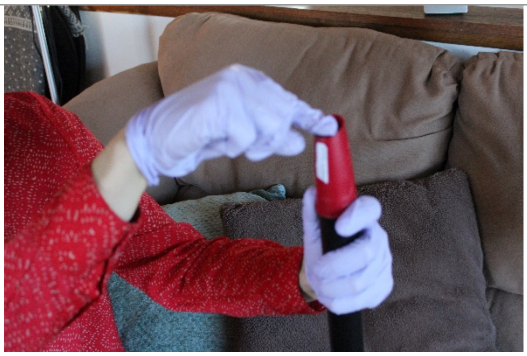

30. Hold the vacuum wand upright in one hand, and place the small black cap on top of the filter and nozzle until it snaps in place

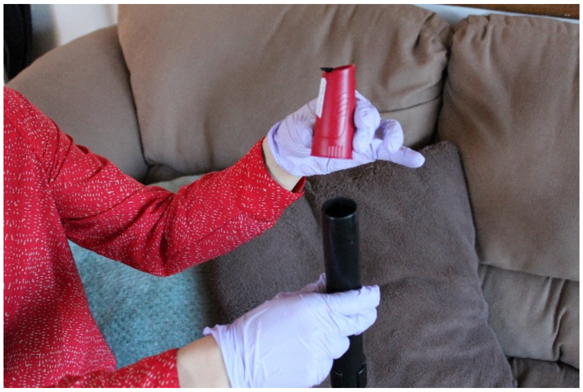

31. Remove the DUSTREAM nozzle from the wand and place the large black cap on the bottom of the nozzle.

<u>Make sure both caps are snapped into place, they are secure and will not come off!</u>

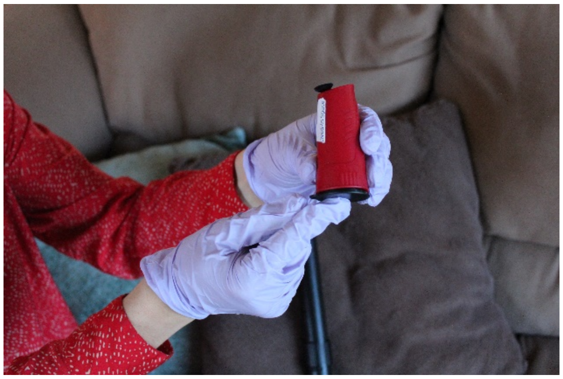

32. Place the entire nozzle into the small ziplock bag it came in.

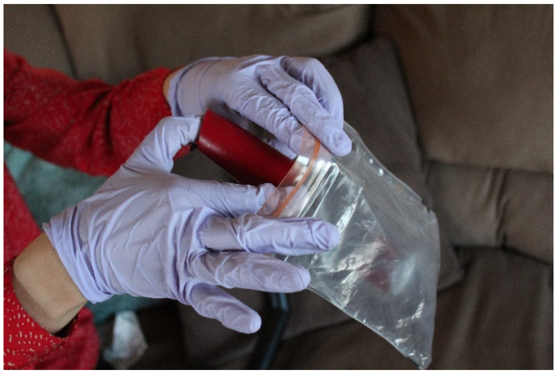

**33. Place both the ziplock bag with the falcon tube and the ziplock bag with the DUSTREAM collector into the dust kit bag they came in and put them in the cooler with ice pack(s).**

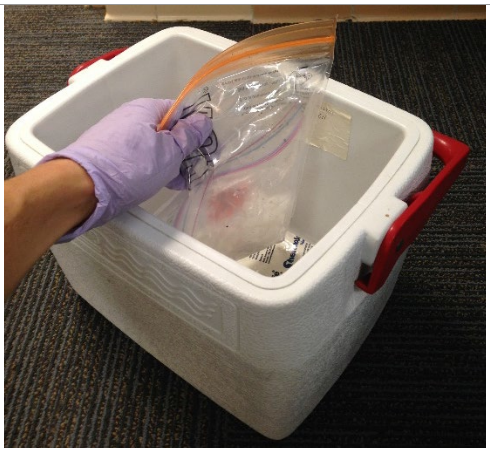

**34. Clean up the sampling site and put all cushions and furniture back to the way it was found.**

**35. Discard of gloves and booties in your garbage bag.**

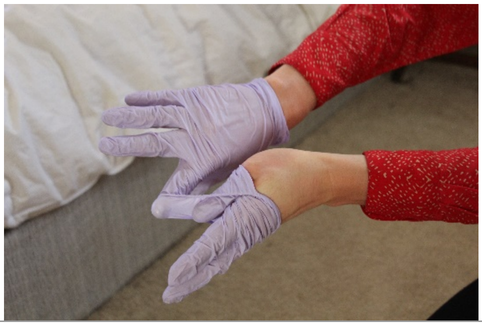
