## Supplementary material for "The Population-based Microbiome Research Core: a longitudinal infrastructure for assessment of household microbiome and human health research": High Touch Surface Swab Collection Protocol

show
survey of the health of wisconsin

School of Medicine and Public Health
UNIVERSITY OF WISCONSIN-MADISON

WISCONSIN MICROBIOME FOLLOW-UP STUDY

### High-Touch Surface Swab Collection Protocol

You will collect a total of 3 swabs, swabbing 4 locations/items per swab*:*

| Kitchen Swab: | Bathroom Swab: | Participant-identified Swab: |
|---|---|---|
| 1. Counter<br>2. Cutting board<br>3. Produce drawer<br>4. Refrigerator handle | 1. Toilet lever<br>2. Sink faucet knobs<br>3. Bathroom sink countertop<br>4. Bathroom door knob | 1. Most-used door knobs (2)<br>2. Most-used device #1<br>3. Most-used device #2 |

Things to remember:

1. Collect 3 swabs total per household
    a. If more than 1 SPID per household, still ONLY collect 3 swabs
    b. Record swabs under 1 SPID in REDcap form, and record in other SPID’s redcap form that samples were collected under other household member
    c. Most-used devices/items include (cellphone, tablet, laptop, remote control, purse, wallet)
2. Swab 4 locations per each swab (as depicted above).
3. Sample each location for 15 seconds (totaling 1 minute per swab).
4. Sample the <u>entire item</u> for the following:
    a. Cutting board (unless larger than sheet of paper)
    b. Refrigerator handle
    c. Toilet lever
    d. Bathroom sink knobs
    e. Door knobs
5. Sample the area of a <u>sheet of paper (8.5 x 11</u>) for the following:
    a. Kitchen counter
    b. Bathroom counter
    c. Cutting board (if large)
6. When sampling electronic devices:
    a. Sample last, to ensure less liquid left on swab
    b. Wipe down with paper towel if wet after swabbing
    c. Wipe back / outside of cell phones & tables, NOT on the front screen

7. If no cutting board/produce drawer/refrigerator handle:
    a. Sample larger counter top area and for longer period of time
        i. Swab sampling time should still be 1 minute

**List of Supplies:**

- Small ziplock bag:
    - 3 packaged swabs
    - 3 labels
        - "Bedroom"
        - "Kitchen"
        - "Other"
- Small ziplock bag:
    - 3 single paper towel sheets
      (wiping surfaces in each location)
- Small ziplock bag:
    - 1 single paper towel sheet
    - "WASTE" label on it
- Disposable non-latex gloves
- Timer
- AAA batteries

### High Touch Surface
### Step-by-Step Guide

#### Overview:

*You will collect a total of three swab samples. There are accompanying questions in REDCap that will help you identify where to swab in the participant's home. Once you have identified the locations, use the following step-by-step guide to collect each of the samples.*

| Instruction | Notes or pictures |
| --- | --- |
| **1. Remove the timer from your sample collection kit and place it somewhere outside of the collection location, but easily within sight.**<br><br>**2. Set timer to 15 seconds** | 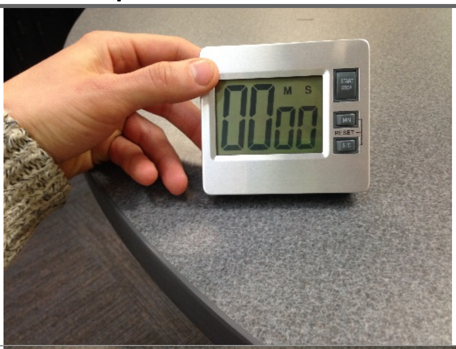 |
| **3. Remove all three swabs from your sample collection kit and set out.** | 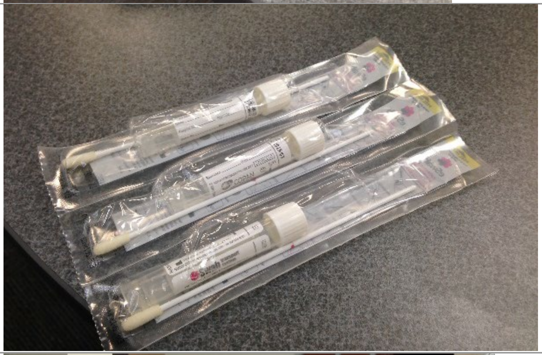 |
| **4. Put on a fresh pair of gloves.** | 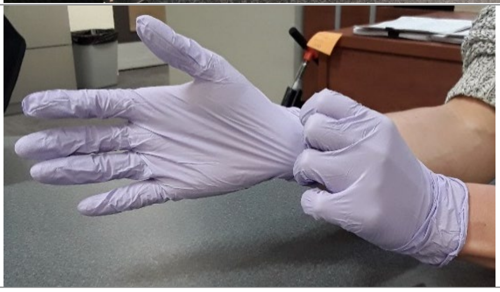 |

| | |
|---|---|
| **4. Take one of the swabs and peel off the plastic wrap <u>ONLY HALF WAY</u>, such that the swab remains in the wrapper. Hold the swab with the wrapper in one hand, and peel back with the other hand.**<br><br>**5. Remove only the tube from the wrapping. Leave swab in wrapper.**<br><br>**6. Unscrew the cap on the tube and place the cap on the counter with the inside facing up toward the ceiling.** | 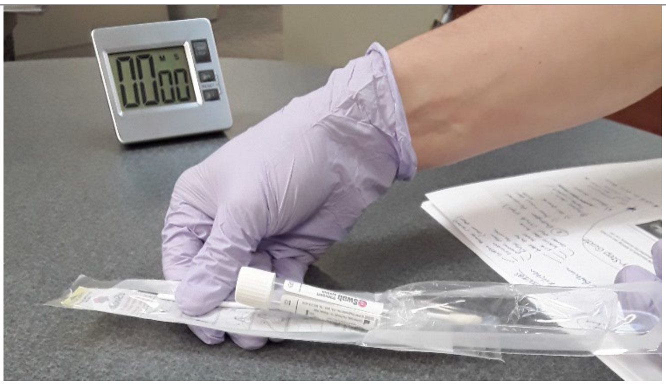 |
| **7. Dip the swab into the tube, fully submerging the swab in the liquid inside the tube.** | 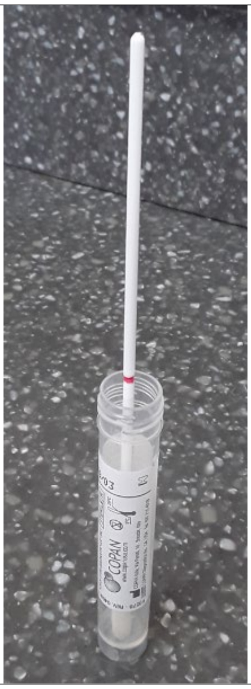 **Swab with liquid in tube** |
| **8. Dump the liquid from the tube into the designated plastic bag in your swab kit.** | |

| | |
|---|---|
| **9. Insert the swab back into the empty swab tube while walking to location where you will swab.** | 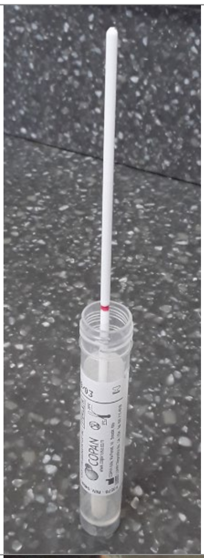 **Swab withOUT liquid in tube** |
| **10. Hit the START button on the timer.**<br><br>**11. Remove the swab from the tube and swab the location for 15 seconds.**<br><br>***ROTATE the swab as you are sampling, making sure all sides of foam tip come into contact with the surface*** | 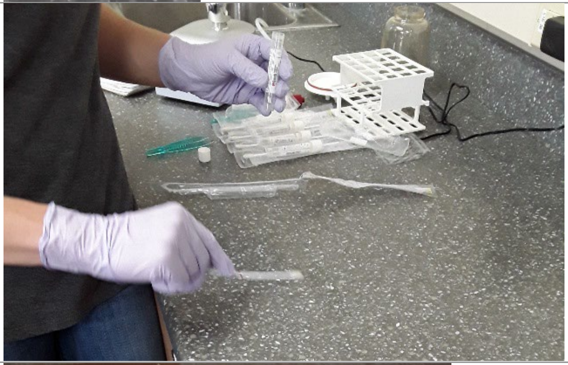 |
| **12. When timer goes off, stop alarm.**<br><br>**13. Place the swab back into the tube, foam tip down so it touches the bottom of the tube.** | 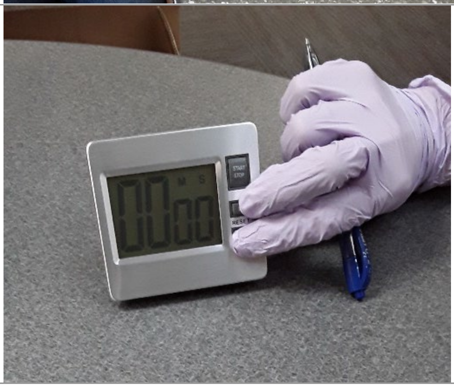 |
| **15. REPEAT steps #8-#10 for the other 3 sampling locations for this swab.** | |
| **16. After you have swabbed all 4 sampling location, put the swab into the tube, with the foam side at the bottom.**<br><br>**The swab will be too long to fit in the tube. With the swab in the tube, break the tube along the red perforation mark.** | 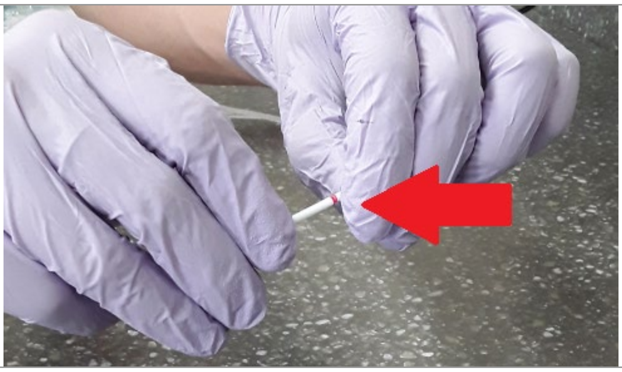 |

| | |
|---|---|
| **17. Discard the end of the swab into your garbage bag. Screw the cap back onto the sample collection tube.** | |
| **18. Put a Brady Barcode SPID label on the tube and the location label on the tube (kitchen, bathroom, or other).**<br><br>**19. Labels should be oriented parallel to the length of the tube and not wrapped around it.** | 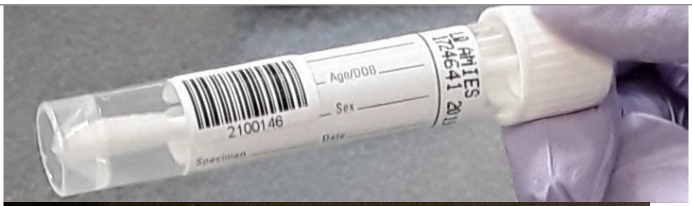  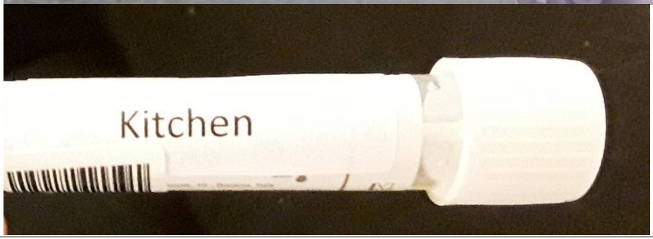  |
| **20. Repeat steps #4-19 for other two swabs.** | |
| **12. Put the sealed tube into the 6x6 ziplock bag it came in and place the bag in the cooler with ices packs after you have collected all three swabs.** | 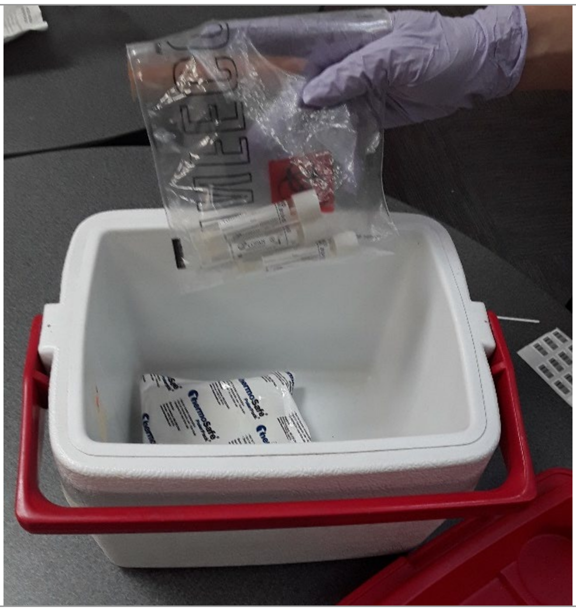 |
