## Supplementary material for "The Population-based Microbiome Research Core: a longitudinal infrastructure for assessment of household microbiome and human health research": Participant Sample Collection Survey

### Dust

The next set of questions ask about your most lived-in space.

Most lived-in space:_____________________________________

**1. When the most lived-in space is cleaned, what type of equipment is used (for example: a vacuum, a broom, a mop, etc.)? <u>Please select all that apply.</u>**

◯ Vacuum
◯ Broom
◯ Mop (including wet mop or Swiffer-type mop)
◯ Don't know
◯ Other → *Specify below.*

**2. Which of the following best characterizes the last time the <u>floor</u> of your most lived-in space was cleaned? <u>Please select one.</u>**

◯ Today
◯ Yesterday
◯ 2-7 days ago
◯ 1-3 weeks ago
◯ More than a month ago
◯ Don't know
◯ Doesn't get cleaned

**3. Which of the following best characterizes the last time <u>furniture, surfaces, and/or walls</u> in your most lived-in space was cleaned? <u>Please select one.</u>**

◯ Today
◯ Yesterday
◯ 2-7 days ago
◯ 1-3 weeks ago
◯ More than a month ago
◯ Don't know
◯ Doesn't get cleaned

### Home

The next set of questions ask about characterisitics of your home.

**1. How many bedrooms does your home have?**

☐☐ bedrooms

**2. Does the home have an open floor plan or rooms that are connected?**

- ◯ Open floor plan (e.g. living room, dining area, and kitchen all in one room)
- ◯ Many rooms connected by doorways
- ◯ Other → *Specify below.*

☐

**3. Which best describes this building? Include all apartments, condos, flats, etc. even if vacant.**

- ◯ A mobile home
- ◯ A one-family house detached from any other house
- ◯ A one-family house attached to one or more houses
- ◯ A building with 2 apartments
- ◯ A building with 3 or 4 apartments
- ◯ A building with 5 to 19 apartments
- ◯ A building with 20 or more apartments
- ◯ A boat, RV, van, etc.
- ◯ Don’t know

STOP

### Soil

The next set of questions ask about your yard or outdoor space.

**1. Is the yard/outdoor space private and only accessible by members of your household or is it shared with others?**

- ○ Shared
- ○ Private
- ○ Don’t Know

**2. Do you or another household member rent or own your home?**

- ○ Own
- ○ Rent
- ○ Don’t Know

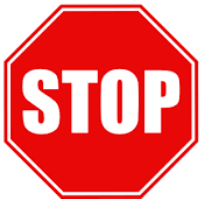

### Surface Swab: Kitchen

The next set of questions ask about your kitchen.

**1. Which of the following best characterizes the last time your kitchen counter was cleaned? <u>Please select one.</u>**

- ◯ Today
- ◯ Yesterday
- ◯ 2-7 days ago
- ◯ 1-3 weeks ago
- ◯ More than a month ago
- ◯ Don’t know
- ◯ Doesn’t get cleaned

**2. What is typically used to clean your kitchen? <u>Please check all that apply.</u>**

- ◯ Soap and water
- ◯ Vinegar
- ◯ Antibacterial spray or wipe (e.g. Lysol or Clorox)
- ◯ Multi-purpose cleaner
- ◯ Don’t know
- ◯ Doesn’t get cleaned
- ◯ Other → *Specify below.*

**3. Does your most-used cutting board get cleaned?**

- ◯ Yes
- ◯ No
- ◯ Don’t Know → **Skip to question 5**
- ◯ Not applicable → **Skip to question 5**

**4. Which of the following best characterizes the last time your most-used cutting board was cleaned? <u>Please select one.</u>**

◯ Today
◯ Yesterday
◯ 2-7 days ago
◯ 1-3 weeks ago
◯ More than a month ago
◯ Don't know
◯ Doesn't get cleaned

**5. Does your refrigerator door handle get cleaned?**

◯ Yes
◯ No
◯ Don't Know → **Skip to question 7**
◯ Not applicable → **Skip to question 7**

**6. Which of the following best characterizes the last time your refrigerator door handle was cleaned? <u>Please select one.</u>**

◯ Today
◯ Yesterday
◯ 2-7 days ago
◯ 1-3 weeks ago
◯ More than a month ago
◯ Don't know
◯ Doesn't get cleaned

**7. Does the produce drawer inside your refrigerator get cleaned?**

◯ Yes
◯ No
◯ Don't Know → **Stop here**
◯ Not applicable → **Stop here**

**8. Which of the following best characterizes the last time your produce drawer was cleaned? <u>Please select one.</u>**

◯ Today
◯ Yesterday
◯ 2-7 days ago
◯ 1-3 weeks ago
◯ More than a month ago
◯ Don't know
◯ Doesn't get cleaned

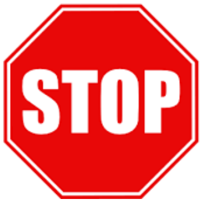

#### Surface Swab: Bathroom

The next set of questions ask about your most frequently used bathroom.

1. **Does your toilet lever get cleaned?**

   ◯ Yes
   ◯ No
   ◯ Don't Know → **Skip to question 3**
   ◯ Not applicable → **Skip to question 3**

2. **Which of the following best characterizes the last time your toilet lever (in your most frequently used bathroom) was cleaned? Please select one.**

   ◯ Today
   ◯ Yesterday
   ◯ 2-7 days ago
   ◯ 1-3 weeks ago
   ◯ More than a month ago
   ◯ Don't know
   ◯ Doesn't get cleaned

3. **What is typically used when your bathroom is cleaned? Please check all that apply.**

   ◯ Soap and water
   ◯ Vinegar
   ◯ Antibacterial spray or wipe (e.g. Lysol or Clorox)
   ◯ Multi-purpose cleaner
   ◯ Don't know
   ◯ Doesn't get cleaned
   ◯ Other → *Specify below.*

   [ ]

4. **Do the knobs on your bathroom sink get cleaned?**

   ◯ Yes
   ◯ No
   ◯ Don't Know → **Skip to question 6**
   ◯ Not applicable → **Skip to question 6**

5. **Which of the following best characterizes the last time the sink knobs (in your most frequently used bathroom) were cleaned? <u>Please select one.</u>**

- ◯ Today
- ◯ Yesterday
- ◯ 2-7 days ago
- ◯ 1-3 weeks ago
- ◯ More than a month ago
- ◯ Don't know
- ◯ Doesn't get cleaned

6. **Does the counter in your bathroom get cleaned?**

- ◯ Yes
- ◯ No
- ◯ Don't Know → **Skip to question 8**
- ◯ Not applicable → **Skip to question 8**

7. **Which of the following best characterizes the last time the bathroom counter (in your most frequently used bathroom) was cleaned? <u>Please select one.</u>**

- ◯ Today
- ◯ Yesterday
- ◯ 2-7 days ago
- ◯ 1-3 weeks ago
- ◯ More than a month ago
- ◯ Don't know
- ◯ Doesn't get cleaned

8. **Does the doorknob inside of your most frequently used bathroom get cleaned?**

- ◯ Yes
- ◯ No
- ◯ Don't Know → **Stop here**
- ◯ Not applicable → **Stop here**

9. **Which of the following best characterizes the last time the inside doorknob (in your most frequently used bathroom) was cleaned? <u>Please select one.</u>**

- ◯ Today
- ◯ Yesterday
- ◯ 2-7 days ago
- ◯ 1-3 weeks ago
- ◯ More than a month ago
- ◯ Don't know
- ◯ Doesn't get cleaned

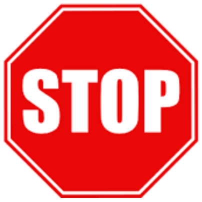

### Surface Swab: Door knobs and items

The next set of questions ask about the following door knobs and items:
**Doorknob #1:**____________________________________________
**Doorknob #2:**____________________________________________
**Surface #1**:_____________________________________________
**Surface #2:**_____________________________________________

**1. Which of the following best characterizes the last time Doorknob #1 was cleaned? <u>Please select one.</u>**

- ◯ Today
- ◯ Yesterday
- ◯ 2-7 days ago
- ◯ 1-3 weeks ago
- ◯ More than a month ago
- ◯ Don't know
- ◯ Doesn't get cleaned

**2. Which of the following best characterizes the last time Doorknob #2 was cleaned? <u>Please select one.</u>**

- ◯ Today
- ◯ Yesterday
- ◯ 2-7 days ago
- ◯ 1-3 weeks ago
- ◯ More than a month ago
- ◯ Don't know
- ◯ Doesn't get cleaned

**3. What is typically used to clean the doorknobs in your house? <u>Please check all that apply.</u>**

- ◯ Soap and water
- ◯ Vinegar
- ◯ Antibacterial spray or wipe (e.g. Lysol or Clorox)
- ◯ Multi-purpose cleaner
- ◯ Don't know
- ◯ Doesn't get cleaned
- ◯ Other → *Specify below.*

**4. Which of the following best characterizes the last time Surface #1 was cleaned? <u>Please select one.</u>**

- ◯ Today
- ◯ Yesterday
- ◯ 2-7 days ago
- ◯ 1-3 weeks ago
- ◯ More than a month ago
- ◯ Don't know
- ◯ Doesn't get cleaned

**5. What is typically used to clean Surface #1? <u>Please check all that apply.</u>**

- ◯ Soap and water
- ◯ Vinegar
- ◯ Antibacterial spray or wipe (e.g. Lysol or Clorox)
- ◯ Multi-purpose cleaner
- ◯ Don't know
- ◯ Doesn't get cleaned
- ◯ Other → *Specify below.*

[ ]

**6. Which of the following best characterizes the last time Surface #2 was cleaned? <u>Please select one.</u>**

- ◯ Today
- ◯ Yesterday
- ◯ 2-7 days ago
- ◯ 1-3 weeks ago
- ◯ More than a month ago
- ◯ Don't know
- ◯ Doesn't get cleaned

**7. What is typically used to clean Surface #2? <u>Please check all that apply.</u>**

- ◯ Soap and water
- ◯ Vinegar
- ◯ Antibacterial spray or wipe (e.g. Lysol or Clorox)
- ◯ Multi-purpose cleaner
- ◯ Don't know
- ◯ Doesn't get cleaned
- ◯ Other → *Specify below.*

[ ]

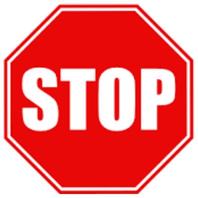
