## Supplementary material for "The Population-based Microbiome Research Core: a longitudinal infrastructure for assessment of household microbiome and human health research": Participant Self-Administered Questionnaire

SPID # ____________ HHID # ____________ SP Initials ____________

### THE WISCONSIN MICROBIOME FOLLOW-UP STUDY QUESTIONNAIRE

**This questionnaire asks you about your health and your home.**

Please complete this booklet on your own with pen or pencil. A SHOW team member will pick it up from you during your sample collection appointment.

**Fill in the bubbles completely.** When you need to write in an answer, please write as clearly as possible in the blanks. You may skip any questions you do not want to answer.

Some of these questions may be familiar; we asked you to answer similar questions the last time we visited. We are asking them again to find out whether anything has changed. There are also some new questions about your health and home that we didn't ask the last time we visited you.

**Thank you for your time in filling out this booklet.** This booklet should take less than one hour to complete.

**If you have any questions, please call 1-888-433-7469.**

show survey of the health of Wisconsin

WISCONSIN MICROBIOME FOLLOW-UP STUDY

School of Medicine and Public Health
UNIVERSITY OF WISCONSIN–MADISON

SHOW – Rev. 2018-01-11

### Your Health History

The questions in this first section ask about your health history. *For each question, please fill in the one circle that describes your health the best or provide a written answer when requested.*

1. **Have you ever had an appendectomy?**

   ◯ Yes
   ◯ No → **Go to Question 3**
   ◯ Don't Know → **Go to Question 3**

2. **How many years ago was your appendectomy?**

   ***For example:***

   | 0 | 7 | years ago

   **Fill in your answer below.**

   | | | years ago

   ◯ Don't know

3. **In the <u>past 12 months</u>, have you had surgery on your digestive system (e.g. esophagus, stomach, liver, appendix, small and large intestines, gall bladder, and/or pancreas)?**

   ◯ Yes
   ◯ No
   ◯ Don't Know

4. **In the <u>past 12 months</u>, have you had any of the following medical devices?**

   ◯ Urinary catheter
   ◯ Vascular catheter
   ◯ Feeding tube
   ◯ Rectal tube
   ◯ Don't know
   ◯ None of the above

**5. In the <u>past 12 months</u>, have you had dialysis treatment?**

- ◯ Yes
- ◯ No
- ◯ Don't Know

**6. In the <u>past 12 months</u>, have you been a patient in a nursing home or inpatient rehabilitation facility?** *An inpatient rehabilitation facility is a facility licensed to provide skilled nursing care and intensive rehabilitation services.*

- ◯ Yes
- ◯ No → **Go to Question 9**
- ◯ Don't Know → **Go to Question 9**

**7. When was your most recent stay in a nursing home or inpatient facility?** *Please tell us what month and year this visit began.*

☐☐ month

☐☐☐☐ year

**8. What was the approximate length of stay?** *Please write the number of days or indicate that you don't know.*

☐☐ days

- ◯ Don't know

**9. In the <u>past 12 months</u>, has a doctor or other health care provider ever told you that you had an infection with a drug-resistant germ?** *A germ is resistant when one or more drugs ordinarily used to treat an infection with that germ cannot kill it.*

- ◯ Yes → *Specify the infection(s) that you had below.*

  ☐

- ◯ No
- ◯ Don't know

**10. In the past 12 months, has a doctor or other health care provider ever told you that you got an infection from a hospital or health care setting?**

- ◯ Yes → *Specify the infection(s) that you had below.*

- ◯ No
- ◯ Don't know

**11. In the past 12 months, have you been put in isolation as a patient in a hospital, nursing home, or inpatient rehabilitation facility?** *That is, visitors were required to wear at least a pair of gloves and gown before seeing you.*

- ◯ Yes
- ◯ No
- ◯ Don't Know

**12.** *Please indicate how much you have been bothered by the following problems over the past 2 weeks.*

**A. Little interest or pleasure in doing things?**

- ◯ Not at all
- ◯ Several days
- ◯ More than half the days
- ◯ Nearly every day
- ◯ Don't know

**B. Feeling down, depressed, or hopeless?**

- ◯ Not at all
- ◯ Several days
- ◯ More than half the days
- ◯ Nearly every day
- ◯ Don't know

#### Household Health History

*These questions ask about the health history of others, rather than yourself.*

**13. In the <u>past 12 months</u>, have you visited someone staying in a healthcare facility (e.g. hosp-ital, nursing home, inpatient rehabilitation facility)?**

○ Yes
○ No → **Go to Question 15**
○ Don't Know → **Go to Question 15**

**14. In the <u>past 12 months</u>, did you provide help in caring for the person(s) while they were staying in the healthcare facility?** *By help in caring, we mean having physical or hands-on contact with the person.*

○ Yes
○ No
○ Don't Know

**15. In the <u>past 12 months</u>, about how many total days did you make a visit to someone who was staying in a healthcare facility (e.g., hospital, nursing home, inpatient rehabilitation facility)?**

[ | ] total number of days

**16. Have you <u>ever</u> visited a person who was placed in isolation while they were in a hospital, nursing home, or inpatient rehabilitation facility?** *That is, you were required to wear at least a pair of gloves and a gown before seeing them.*

○ Yes
○ No
○ Don't Know

**17. Has anyone in your household <u>ever</u> had an infection with a drug-resistant germ?** *A germ is resistant when one or more drugs ordinarily used to treat an infection with that germ cannot kill it.*

○ Yes
○ No
○ Don't Know

**18. Has anyone in your household <u>ever</u> had an infection from a hospital or healthcare setting?**

- ○ Yes → *Specify the infection(s) that they had below.*

  [ ]

- ○ No
- ○ Don't know

#### Prescription Medications and Select OTC Drugs

The next set of questions will gather information about any medication you might be taking. *For each question, please fill in the one circle that comes closest to your current medication use, or fill in the blanks as requested.*

**19. In the past 3 months, have you taken an antibiotic (a drug used to treat an infection)?**

- ○ Yes
- ○ No → **Go to Question 22**
- ○ Don't Know → **Go to Question 22**

**20. In the table below, please list the name(s) of the antibiotics, the illness or condition for which you took them, and the length of time you took them.** *If you were prescribed the same antibiotic more than once in the past year, list it multiple times. Feel free to get your medication bottles if you need to reference the medication name.*

| *Name the antibiotic you took in the last year below:* | *The reason (illness or condition) for taking the medication:* | *For how many days did you take this antibiotic?* |
|---|---|---|
| *1.* | | |
| *2.* | | |
| *3.* | | |
| *4.* | | |
| *5.* | | |

☐ **If you have had <u>more than five</u> antibiotic prescriptions in the past year, please check this box.**

**21. When was the last time you took any antibiotics?**

○ Today **or** ☐☐ number of days ago

22. **Are you currently using probiotic supplements?** *Specifically, we are referring to pills containing healthy bacteria.*

- ◯ Yes
- ◯ No → **Go to Question 24**
- ◯ Don't Know → **Go to Question 24**

23. **When was the last time you took the probiotic supplement(s)?**

◯ Today **or** [ | ] number of days ago

24. **In the past 12 months, have you taken a proton pump inhibitor? Proton pump inhibitors are drugs that suppress the production of acid in your stomach.** *Some examples of trade (generic) names are: Aciphex (rabeprazole), Protonix (pantoprazole), Nexium (esomeprazole), Prevacid (lansoprazole), Kapidex (dexlansoprazole), Zegerid (omeprazole/sodium bicarbonate), Prilosec (omeprazole), Dexilant (dexlansoprazole).*

- ◯ Yes
- ◯ No → **Go to Question 26, page 8**
- ◯ Don't Know → **Go to Question 26, page 8**

25. **Are you currently taking a proton pump inhibitor?**

- ◯ Yes
- ◯ No
- ◯ Don't Know

#### Smoking and Other Tobacco Products

The next questions are about your history of using tobacco products. *For each question, please fill in the circle that comes closest to your current use of tobacco products, or fill in the blank as requested.*

**26. Have you smoked 100 or more cigarettes in your entire life?**

○ Yes ○ No → **Go to question 30**

**27. How old were you when you started smoking cigarettes regularly?**

Enter age when you started smoking: [ ][ ]

**28. Do you smoke cigarettes now?**

○ Yes ○ No → **Go to question 30**

**29. On average, when you smoked during the past 30 days, about how many cigarettes did you smoke per day?** *If you smoked less than 1 cigarette per day, enter 1 (1 pack = 20 cigarettes).*

Enter number of cigarettes per day: [ ][ ]

**30. How many hours per week are you currently exposed to cigarette smoke in social settings outside your own home? (This would include time spent with friends or relatives who smoke, time spent in restaurants or taverns, or other social affairs where people are smoking.)**

***For example:***

[0][0][7] hours per week

**Fill in your answer below.**

[ ][ ][ ] hours per week

○ Don't know

**31. Do any people currently smoke cigarettes inside your home?**

- ◯ Yes
- ◯ No
- ◯ Don’t Know

**32. Do you now smoke tobacco products other than cigarettes every day, some days, or not at all?**

- ◯ Every day
- ◯ Some days
- ◯ Not at all

**33. Do you now use any smokeless tobacco products, such as chewing tobacco, snuff, snus, dip, orbs, sticks, or strips?**

- ◯ Every day
- ◯ Some days
- ◯ Not at all

**34. Do you now use electronic cigarettes (e-cigarettes) every day, some days, or not at all?**

- ◯ Every day
- ◯ Some days
- ◯ Not at all

**35. Which of the following would you say comes closest to what happens inside your home?**

- ◯ Smoking is not allowed inside my home
- ◯ Smoking is allowed everywhere
- ◯ Smoking is allowed only in certain areas inside my home
- ◯ Smoking is allowed only for special guests inside my home
- ◯ Have not thought about it
- ◯ Other, specify: ____________________

#### Housing Characteristics

The next section asks questions about your home and your exposure to certain hazards in the home. *For each question, please fill in the one circle that comes closest to describing the qualities of your home.*

**36. Have you done any improvements, renovations or extensions to your home in the past three months?**

◯ Yes
◯ No
◯ Don't Know

**37. On average, how often do you open the windows in your home for longer than an hour during <u>mid-winter</u>?**

◯ Never
◯ 1-2 times per month
◯ 1-2 times per week
◯ More than twice per week

**38. On average, how often do you open the windows in your home for longer than an hour during <u>mid-summer</u>?**

◯ Never
◯ 1-2 times per month
◯ 1-2 times per week
◯ More than twice per week

**39. Have you used a dehumidifier in your home in the past <u>12 months</u>?**

◯ Yes
◯ No → **Go to Question 42 page 11**
◯ Don't Know → **Go to Question 42, page 11**

**40. If yes, how often on average do you use a dehumidifier?**

◯ Rarely
◯ Occasionally
◯ Regularly
◯ Constantly

**41. If yes, do you use a dehumidifier seasonally (e.g. only in the summer), or throughout the year?**

- ◯ Seasonally
- ◯ Year-Round
- ◯ Don't Know

**42. Are you aware of any leaks or water problems in your home in the <u>past three months</u>?**

- ◯ Yes
- ◯ No → **Go to Question 44**
- ◯ Don't Know → **Go to Question 44**

**43. What type of leak was it?**

- ◯ Roof leak
- ◯ Basement leak
- ◯ Plumbing
- ◯ Sewer back up
- ◯ Window leak
- ◯ Flood
- ◯ Overhead apartment leak
- ◯ Toilet overflow
- ◯ Other, specify: ______

**44. What type of cooking appliance(s) do you have in your home? (select all that apply)**

- ◯ Gas or propane stove
- ◯ Electric stove
- ◯ Wood burning stove
- ◯ None
- ◯ Don't know
- ◯ Other, specify: ______

**45. What type of air conditioning does your residence have?**

- ◯ Don't have A/C → **Go to Question 47, page 12**
- ◯ Central A/C
- ◯ Window, wall of portable unit(s)
- ◯ Don't Know → **Go to Question 47, page 12**

**46. How often do you use your air conditioning <u>in the summer</u>?**

- ◯ Never
- ◯ Rarely
- ◯ Occasionally (average once or more per week)
- ◯ Regularly (turned on most of the time with windows kept closed)
- ◯ Don't know

**47. In the past <u>12 months</u>, have you used a portable air cleaner/purifier in your home?**

- ◯ Yes
- ◯ No → **Go to Question 49**
- ◯ Don't Know → **Go to Question 49**

**48. How often do you use your portable air cleaner/purifier?**

- ◯ Never
- ◯ Rarely
- ◯ Occasionally
- ◯ Regularly
- ◯ Don't know

**49. Are there members of the household who work with hazardous materials on the job?**

- ◯ Yes, please specify: ________
- ◯ No
- ◯ Don't Know

**50. Is your home within 100 yards of any of the following?**

- ◯ Major Highway/Artery
- ◯ Factory
- ◯ Gas station
- ◯ Body of water
- ◯ Farm
- ◯ Large parking lot
- ◯ Major/prolonged construction activity (e.g. building of houses or other buildings, road work, typically involving heavy machinery and/or generation of noticeable amounts of dust in the air)
- ◯ None of these choices
- ◯ Other source of pollution, please specify: ________

#### Where You Spend Your Time

This next set of questions will ask you about where you typically spend time. *Please fill in the blank next to each item, indicating where you typically spend your time.*

**51. How much time (in hours) per day/night do you spend in the following locations?**

**a. On a typical WEEKEND day in the SUMMER months (May through October)**

*Hours must total 24 for each question. Use whole numbers only; enter 00 if no time spent.*

| Hours | Location |
|---|---|
| | **Home indoors (including sleeping)** |
| | **Outdoors near home (in yard, local park, walk/bike in neighbourhood)** |
| | **On roadways (car/van, bus)** |
| | **Other forms of transit (train, subway)** |
| | **Other indoor places** |
| | **Other outdoor places** |
| 24 | **TOTAL** |

**b. On a typical WEEK day in the SUMMER months (May through October)**

*Hours must total 24 for each question. Use whole numbers only; enter 00 if no time spent.*

| Hours | Location |
|---|---|
| | **Home indoors (including sleeping)** |
| | **Outdoors near home (in yard, local park, walk/bike in neighbourhood)** |
| | **On roadways (car/van, bus)** |
| | **Other forms of transit (train, subway)** |
| | **Other indoor places** |
| | **Other outdoor places** |
| 24 | **TOTAL** |

**c. On a typical WEEKEND day in the WINTER months (November through April)**

*Hours must total 24 for each question. Use whole numbers only; enter 00 if no time spent.*

| Hours | |
|---|---|
| ☐☐ | **Home indoors (including sleeping)** |
| ☐☐ | **Outdoors near home (in yard, local park, walk/bike in neighbourhood)** |
| ☐☐ | **On roadways (car/van, bus)** |
| ☐☐ | **Other forms of transit (train, subway)** |
| ☐☐ | **Other indoor places** |
| ☐☐ | **Other outdoor places** |
| 2 4 | **TOTAL** |

**d. On a typical WEEKEND day in the WINTER months (November through April)**

*Hours must total 24 for each question. Use whole numbers only; enter 00 if no time spent.*

| Hours | |
|---|---|
| ☐☐ | **Home indoors (including sleeping)** |
| ☐☐ | **Outdoors near home (in yard, local park, walk/bike in neighbourhood)** |
| ☐☐ | **On roadways (car/van, bus)** |
| ☐☐ | **Other forms of transit (train, subway)** |
| ☐☐ | **Other indoor places** |
| ☐☐ | **Other outdoor places** |
| 2 4 | **TOTAL** |

#### Home Cleaning, Products and Appliances

These next questions ask about the household cleaning habits and products used in and around your household. *For each question, please fill in the one circle that comes the closest to describing what happens in your home.*

**52. When people come into your home, do they usually remove their shoes?**

- ◯ Yes
- ◯ No
- ◯ Don't Know

**53. Is a vacuum cleaner ever used to clean the floors in your home?**

- ◯ Yes
- ◯ No → **Go to Question 56, page 16**
- ◯ Don't Know → **Go to Question 56, page 16**

**54. How frequently are the floors in the main living space vacuumed?**

- ◯ Daily
- ◯ Weekly
- ◯ Monthly
- ◯ Less than monthly
- ◯ Never
- ◯ Don't know

**55. How frequently is the floor in your bedroom vacuumed?**

- ◯ Daily
- ◯ Weekly
- ◯ Monthly
- ◯ Less than monthly
- ◯ Never
- ◯ Don't know

**56. During the last <u>12 months</u>, how often were weed killers or insecticides used on the foundation, yard/lawn, flowers, vegetables or fruit trees outside your house?**

- ○ 0 times
- ○ 1 time
- ○ 2 - 3 times
- ○ 4 - 6 times
- ○ > 10 times
- ○ Don't know

**57. During the last <u>12 months,</u> how often were chemicals such as pesticides used inside your home to kill or control insects or other pests?**

- ○ 0 times → **Go to question 59, page 17**
- ○ 1 time
- ○ 2 - 3 times
- ○ 4 - 6 times
- ○ > 10 times
- ○ Don't know → **Go to question 59, page 17**

**58. Which rooms in your home were treated with this product? Fill in all that apply**

- ○ Kitchen
- ○ Bathroom
- ○ Living room or family room
- ○ Bed
- ○ Laundry
- ○ Basement
- ○ Don't know
- ○ Other, please specify: ____________

**59. Select all of the following statements with which you agree:**
***Cleaning around the house is something ...***

- ◯ I do frequently
- ◯ I do automatically
- ◯ I do without having to consciously remember
- ◯ That makes me feel weird if I do not do it
- ◯ I do without thinking
- ◯ That would require effort not to do it
- ◯ That belongs to my (daily, weekly, monthly) routine
- ◯ I start doing before I realize I'm doing it
- ◯ I would find hard not to do
- ◯ I have no need to think about doing
- ◯ That's typically "me"
- ◯ I have been doing for a long time
- ◯ None of the above

#### Household Water Use

The following questions ask about the water use in your home. *For each question, please fill in the circle next to the option that comes closest to describing the water use in your home.*

**60. Do you use a water filter or treatment system such as an aerator, Brita filter, carbon filter, water softener, refrigeration filtration, distillation, reverse osmosis, or other filtration treatment system in your home for drinking water?**

- ◯ Yes
- ◯ No → **Go to Question 62, page 19**
- ◯ Don't Know → **Go to Question 62, page 19**

**61. Please select all that apply regarding the filters or treatments that you use on your drinking water.**

- ◯ Pitcher type water filter
- ◯ Refrigeration filtration system
- ◯ Treat at point of use/under the sink. *Indicate which types.*
  - ◯ Carbon filter
  - ◯ Reverse osmosis
  - ◯ Water softener
  - ◯ Distillation
  - ◯ Absorbent media (iron-oxide filter)
  - ◯ Don't know
- ◯ Treat all water in the home. *Indicate which types.*
  - ◯ Carbon filter
  - ◯ Reverse osmosis
  - ◯ Water softener
  - ◯ Distillation
  - ◯ Absorbent media (iron-oxide filter)
  - ◯ Don't know
- ◯ Drink only purchased bottled water
- ◯ Don't know
- ◯ Other, please specify: ____________________

**62. In a typical day, how many 8 oz. servings of tap water do you drink at home? (an 8 oz. serving equals one cup, a ¼ liter, or ¼ of a quart)**

[ | ] servings

**63. What type of water do you typically drink or use to prepare drinks at home?**

- ○ Plain tap
- ○ Filtered
- ○ Softened
- ○ Bottled
- ○ Don't know

**64. About how many <u>showers</u> do you take during a TYPICAL WEEK?** *Enter 00 if you do not take showers.*

***For example:***

[ 0 | 7 ] showers

**Fill in your answer below.**

[ | ] showers

○ Don't know

**65. About how many <u>baths</u> do you take during a TYPICAL WEEK?** *Enter 00 if you do not take baths.*

***For example:***

[ 0 | 7 ] baths

**Fill in your answer below.**

[ | ] baths

○ Don't know

**66. On average, how long is a typical <u>shower</u>?** *Enter 00 if you do not take showers.*

***For example:***

| 0 | 8 |
|---|---|

minutes

**Fill in your answer below.**

minutes

○ Don't know

**67. On average, how long is a typical <u>bath</u>?** *Enter 00 if you do not take baths.*

***For example:***

| 0 | 8 |
|---|---|

minutes

**Fill in your answer below.**

minutes

○ Don't know
