## Supplementary material for "The Population-based Microbiome Research Core: a longitudinal infrastructure for assessment of household microbiome and human health research": Soil Collection Protocol

#### Overview:

- You will collect soil from 2-3 locations in the yard / outdoor space surrounding the home.
- One 1-mL cryovials of soil will be collected from each of the 2-3 locations
- This totals 2-3 soil samples (1 mL each) per household

**How to determine where to sample:**

| | |
|---|---|
| Sample location 1: | Near exit/entrance of the home used most often by participant & other household members |
| Sample location 2: | Participant-identified locations where participant and other household members spend the most time and/or frequent the most |
| Sample location 3 (Optional): | Participant's garden. Only sample from a garden if the participant reports spending time in their garden. |

**Near exit / entrance sample:**

- Select soil from a location as close to the entrance/exit as possible
- For apartment buildings, sample nearest to the exit /entrance of the building door or parking garage used

**Participant-identified location:**

- Participants think about all their outdoor activities, chores and habits and chose 1 location where they and other household members spend the most time

  ***Participant will use a handcard to prompt them to think about how they spend time in their yard:***

  - Examples of activities participants are prompted to consider are:
    - Sitting on the patio, grilling, gardening, watering plants, construction projects, dining, socializing, playing/recreating, mowing the lawn, working on the car, etc.
  - Participants are also asked to think about possible chores that they do around the yard such as:
    - Getting the mail, taking the trash out

- If participant DOES NOT spend time in outdoor spaces for more than 5 minutes or DOES NOT have any outdoor spaces

- Sample along pathway where participant/household members do chores:
    - Walk to car, take out trash, get mail, etc.
    - Sample near or under a commonly used window/patio/balcony

**List of Supplies:**

a. Soil Collection Kit:
    i. Small ziplock bag:
        1. (3) 1mL cryovials
        2. Soil ID labels
            a. (1) with #1 label
            b. (1) with #2 label
            c. (1) with #3 label
b. Soil corer
c. Bottle brush
d. Spray bottle filled with water
e. Metal spatula and/or scoopula
f. Non-latex gloves

### Soil Step-by-Step Guide

#### Overview:

*You will collect soil from five different locations in participant's yards. Accompanying questions in REDCap will help you identify those locations. Once you have determined where you will sample, reference this step-by-step guide to collect the samples.*

| | |
|---|---|
| **1. Identify where you will take the first soil core sample** | *You will identify this by asking participant questions in REDcap.* |
| **2. Put on a fresh pair of gloves.** | 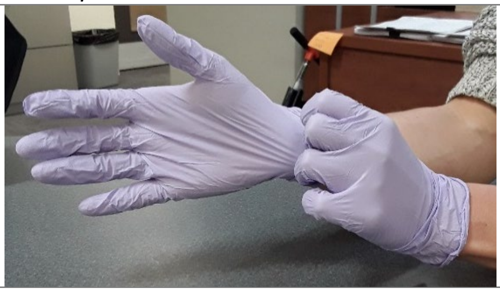 |
| **3. At this first location, take the soil corer and push it into the ground – as deep as you can manage (at least 10 inches)– and then pull it out.** | *Step 3 & 4 help to "cleanse" the soil corer and remove any dirt from a prior household.* 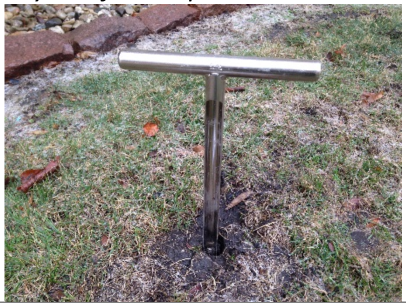 |
| **4. Remove all soil from the soil corer** | *May need to use metal scoopula or bottlebrush to do so.* 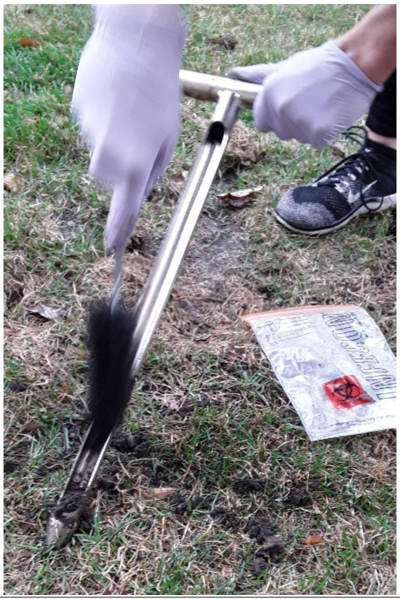 |

**5. Repeat step #3, pushing the soil corer into the ground near where you just cored.**

**6. Place the soil corer on the ground with the open window slot facing up toward the sky.**

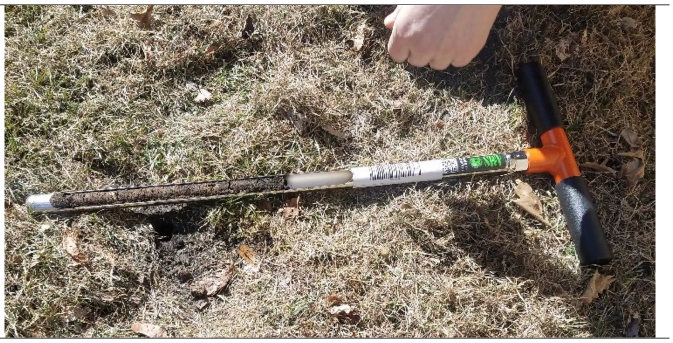

**7. Take ziplock bag of 1mL tubes out of the soil collection kit bag.**

**8. Place the empty soil kit bag flat on the ground.**

*This will be used to place tube caps on upside down when filling tube with soil.*

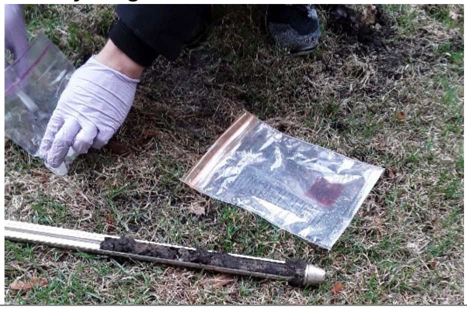

**9. Take one 1mL tube out of the ziplock bag that has the correct # SOIL LABEL on it that matches the soil number # in REDcap.**

*Your tubes should already be labeled with the SPID and a number corresponding to the sample documentation in REDCap. Be sure that you are using the correct tubes.*

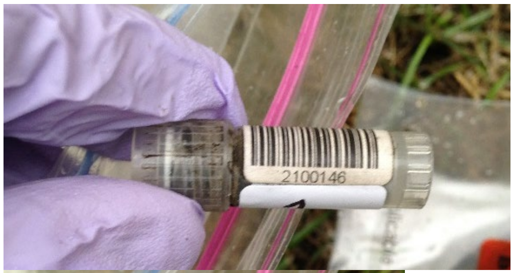


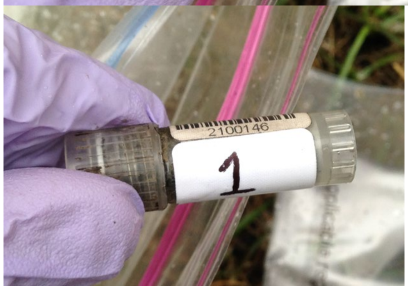

| | |
|---|---|
| **Unscrew the tube and place the cap upside down (open end facing sky) on top of the soil bag.** | 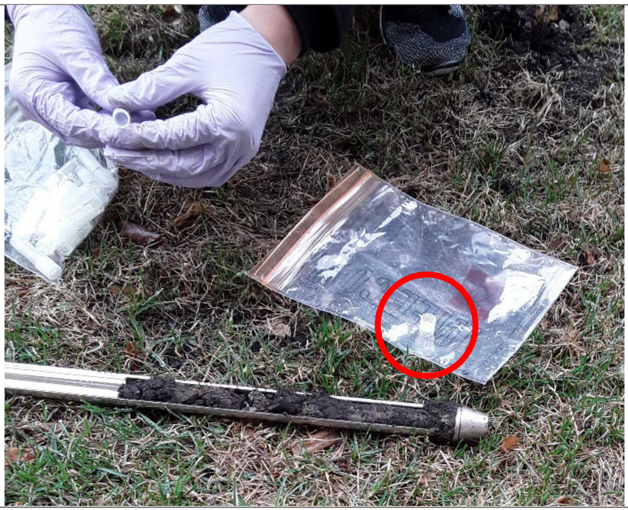 |
| **10. Fill up the 1ml tube with soil, taking soil from nearest the bottom of the soil corer (farthest from the handle side).**<br><br>**11. Use metal scoopula to help get the soil into the cryovial.**<br><br>**12. Use a metal spatula for compact soil to poke / press the soil all the way to the bottom of the cryovial.** | *Do not take soil from the top layer of the sample – make sure you take soil deep enough that you do not include any roots from grass or other plants, or from the surface soil.*<br>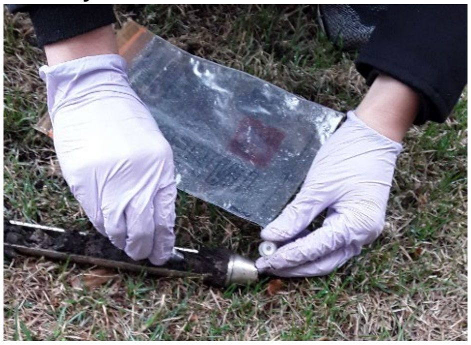<br>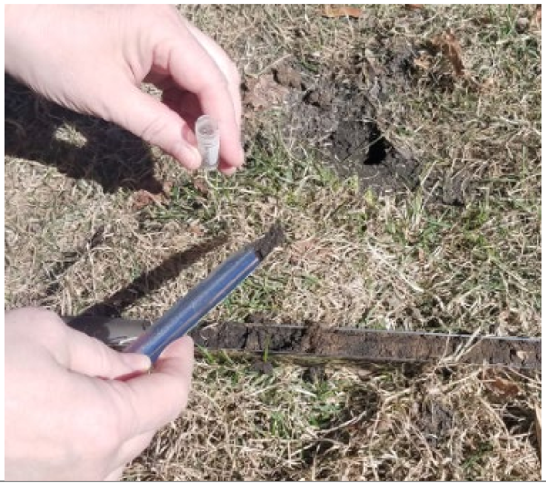 |

| | |
|---|---|
| **11. After the tube is full of soil, screw the cap back on and place back in the ziplock bag it came from.** | *Your tubes should already be labeled with the SPID and a number corresponding to the sample documentation in REDCap. Be sure that you are using the correct tubes.* 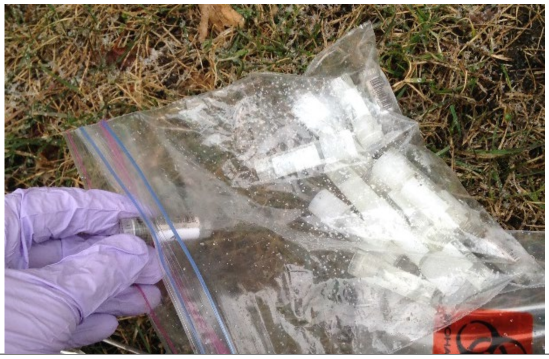 |
| **12. After collecting the soil sample from this location, clean soil out of corer.** | *Shake soil out and/or use bottle brush. Do NOT clean with water at this point.* 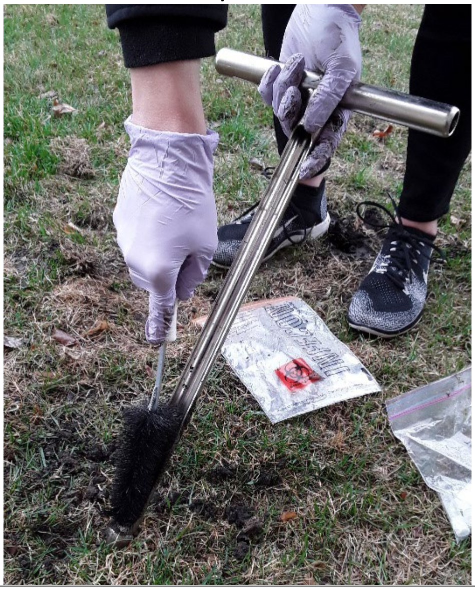 |
| **13. Do your best to put any ground cover, mulch or top soil back over the soil core hole.** | |
| **15. Repeat steps #5-13 until you have cored a maximum of 3 locations and collected a maximum of 3 1mL tubes of soil.** | *Ensure you are using the correct tubes with the correct soil ID number on them – they need to match what is in REDcap.* |
| **16. When you have finished soil collection, place the 6x6 ziplock bag with the 1mL tubes into you cooler with ice pack(s).** | *If an extra / unused tubes, keep with your belongings and SHOW will collect from you.* |

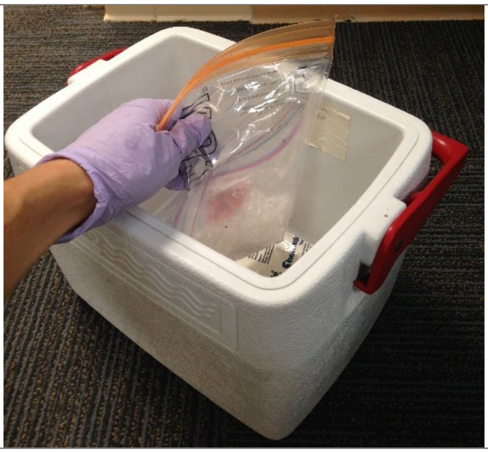

**Between household visits be sure to clean soil corer out with water.**

***Use spray bottle given, or other method in your home (outdoor hose).***

***DO NOT USE CLEANING PRODUCT ON THE SOIL CORER! WATER ONLY!***
