## Supplementary material for "The Population-based Microbiome Research Core: a longitudinal infrastructure for assessment of household microbiome and human health research": Stool Collection Instructions for Participant

### Instructions for Stool Collection

*Thank you for donating this important sample.*

#### 1. Please wash hands before beginning procedure.

COLLECT stool within 24 hours of your home visit appointment (not any earlier)
PLEASE DO NOT pass the stool directly into the toilet.
PLEASE DO NOT pass the stool directly into the collection vial.
PLEASE DO NOT urinate on the stool sample.

#### 2. Place the tub and frame into the toilet to collect the stool sample.

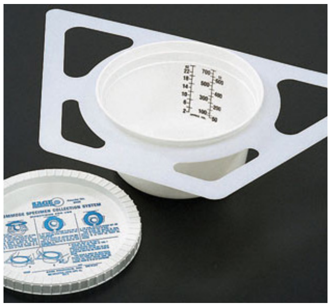

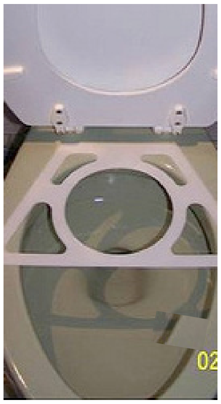

- Remove the lid from the plastic collection tub or 'margarine tub', and put the tub through the round hole of the trapezoid-shaped frame.
- Lift the toilet seat and place the tub and frame along the back edge of the toilet, so the long straight side is facing the center of the toilet.
- Sit on the toilet as usual and have the bowel movement into the tub.

#### 3. Put on gloves (provided) and use the wooden tongue depressor (shown in picture) to scoop out several tablespoons of stool and put this into the sterile plastic container (shown in picture).

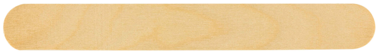

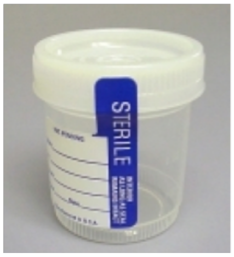


- Close the top of the sterile container tightly to avoid leakage.
- <u>Record date and time of collection on outside of container</u>
- Discard used 'margarine tub' with the lid on in the regular trash.

#### 4. Place the container in the biohazard bag, seal bag tightly. Then place into paper bag.

WISCONSIN MICROBIOME FOLLOW-UP STUDY

#### 5. Store the sample in your refrigerator until your home visit appointment when a SHOW field staff member will collect the stool.

**If you have any problems or questions, please call SHOW at 888-433-7469.**

show survey of the health of Wisconsin
